# Third dose COVID-19 vaccination elicits immune memory in patients with inborn errors of immunity even in absence of neutralizing antibodies

**DOI:** 10.1101/2025.11.09.25339598

**Authors:** Emily S.J. Edwards, Raffi Gugasyan, Nirupama Varese, Shir Sun, Jessica E. Canning, Ebony G. Blight, Pei M. Aui, Irene Boo, Stuart Turville, Anupriya Aggarwal, Samar Ojaimi, Julian J. Bosco, Stephanie Stojanovic, P. Mark Hogarth, Heidi E. Drummer, Scott J. Bornheimer, Robyn E. O’Hehir, Menno C. van Zelm

**Author notes:** Authors contributed equally (Emily S.J. Edwards, Raffi Gugasyan). **Correspondence:** Emily S.J. Edwards, PhD, Allergy and Clinical Immunology Laboratory, Monash University, 89 Commercial Rd, Melbourne, VIC 3004, Australia., Menno C. van Zelm, PhD, Department of Immunology, Erasmus MC, University Medical Center, Dr. Molewaterplein 40, 3015 GD Rotterdam, the Netherlands.

## Abstract

**Background:** Immunocompromised people, including those with Inborn Errors of Immunity (IEI), are at increased risk of severe disease from viral infections. Therefore, regular booster vaccinations are recommended for SARS-CoV-2 and influenza, but it is unclear if these elicit protective immunity.

**Objective:** Comprehensive evaluation of adaptive immune responses, including SARS-COV-2 specific antibodies, memory B-(Bmem) and memory T-cells (Tmem), to COVID-19 vaccination in IEI patients.

**Methods:** Blood samples were collected at 1-month post doses 2 and 3 of the ancestral COVID-19 vaccine, SARS-CoV-2 neutralizing antibodies (NAb) and Spike receptor binding domain (RBD) specific IgG were determined in 25 IEI patients and 29 controls. Ancestral Spike specific Tmem, and ancestral and Omicron subvariant RBD-specific Bmem were evaluated with flow cytometry.

**Results:** After dose 2, IEI patients had significantly lower Nab, RBD-specific IgG and Bmem against ancestral and Omicron subvariants. Third dose vaccination boosted NAb, IgG and Bmem levels, but these remained lower than healthy controls. Especially IgG1^+^ Bmem were lower in the IEI patients, while they carried higher frequencies of CD71^+^ ancestral RBD-specific Bmem. IEI patients and controls had similar numbers of Spike-specific CD4^+^ and CD8^+^ Tmem after both doses. However, patients’ Tmem had lower CD69 expression and reduced cytokine co-expression. While 9/25 IEI patients did not have NAb after dose 3, all had detectable SARS-CoV-2 specific IgG, Bmem- and/or Tmem.

**Conclusion:** Patients with IEI form lower levels of antibodies and immune memory cells to COVID-19 vaccination than controls. Still, all patients displayed formation of adaptive immune memory. This suggests a beneficial effect of vaccination, and supports the strategy for offering regular booster vaccinations to limit severe COVID-19 in this at-risk population.

**KEY MESSAGES:**

- Whilst antibody-deficient patients fail to generate NAb, all evaluated IEI patients could form spike-specific Tmem after COVID-19 vaccination.
- A 3-dose regimen boosted humoral and/or cellular responses in 60% of IEI patients, reinforcing the importance of prioritizing this population for early and repeated vaccination.
- Comprehensive analysis of Bmem and Tmem responses revealed COVID-19 vaccination elicited adaptive immunity, underscoring the need for monitoring of immune cells in addition to serology in immunodeficient populations.

**CAPSULE SUMMARY:** Following 3-dose COVID-19 vaccination, all adult patients with inborn errors of immunity formed memory B- and/or memory T cells regardless of whether neutralizing antibodies are elicited. Therefore, IEI patients generate adaptive immunity to COVID-19 vaccination.

## INTRODUCTION

Inborn Errors of Immunity (IEI) are rare genetic disorders that impair immune function, leading to increased susceptibility to infections, as well as chronic lung disease, autoimmunity, gastrointestinal disease and cancer. Previously named primary immunodeficiencies, IEI now include disorders that lead to immune dysregulation and can be caused by variants in more than 508 genes.^1, 2^

Patients with IEI are at heightened risk for severe COVID-19 infections,^3–5^ with studies reporting a three- to four-fold higher risk of hospitalization, five-fold higher risk of intensive care unit admission,^5^ and eight-fold higher case fatality rate compared to the general population.^3, 5, 6^ This vulnerability is attributed to defects in antibody production, T-cell function and/or innate immune responses, which collectively compromise viral clearance and immune protection.^1, 7–9^

In 2021, healthcare workers and immunocompromised patients including those with IEIs were prioritized for access to COVID-19 vaccination in Australia.^10, 11^ The primary vaccine regimen for healthy adults was two doses of mRNA (Pfizer BioNTech BNT162b2) or adenoviral (ChAdOx1) vaccines, while immunocompromised groups including those with IEIs, were given a third dose (mRNA either Pfizer BioNTech BNT162b2 or Moderna mRNA1273) to boost immunity and reduce the risk of infection and/or severe COVID-19 disease.^10–13^ In Australia, due to border closures and COVID-19 restrictions, SARS-CoV-2 infection rates remained low until July 2021, when the Delta variant caused a surge in infections. Nevertheless, overall infection rates remained low, allowing most adults to receive three vaccine doses prior to the large case wave by Omicron BA.1 and subsequent Omicron sub-lineage emergence.^14–18^

To combat COVID-19, new vaccine platforms were utilized, and included the mRNA and adenoviral platforms, which were both shown to be effective in prevention of severe disease in the general population.^19–21^ This was associated with the capacity to elicit a neutralizing antibody response,^14, 16, 22–24^ and the formation of long-lived Bmem ^14–16, 25, 26^ and Tmem.^26–32^ Importantly, the primary output for vaccine trials and the gold standard for assessment of vaccine responses in the clinical setting is quantification of NAb.^33^ To date, studies have shown that seroconversion rates are lowest in patients with common variable immunodeficiency (CVID) or combined humoral and cellular immunodeficiency (CID), whereas patients with clinically milder antibody deficiencies, including IgA deficiency and specific polysaccharide antibody deficiency (SpAD), display responses more similar to healthy controls.^34–40^ Complicating antibody assessment further, many IEI patients receive immunoglobulin replacement therapy (IgRT), which prevents distinction of patient-derived antibody responses from antibodies obtained from supplementation therapy, thus preventing accurate measurement of vaccine induced antibodies in this population. As antibody responses are likely not an accurate measure of vaccine effectiveness in IEI patients,^41^ it is important to determine the efficacy of COVID-19 vaccination in eliciting robust Bmem and Tmem immunity, to prevent and/or limit severe COVID-19 infection.

Reports of T-cell responses in IEI patients showed varied outcomes from largely normal in those with X-linked agammaglobulinemia,^39, 42–45^ to significantly reduced responses in CVID and CID.^34, 35, 46–49^ Currently, studies on Bmem responses to COVID-19 vaccination in IEI are limited to detection of IgG positive B cells via ELISPOT.^36, 50, 51^ This leaves a gap in understanding about the quantity, phenotype and variant-binding capacity of spike-specific Bmem induced by COVID-19 vaccination in IEI patients. In this study, we provide a comprehensive analysis of the adaptive immune response to COVID-19 vaccination in adults with IEI, encompassing NAb titers, specific IgG, Bmem and Tmem. Importantly, those patients who failed to mount NAb titers, generated Tmem and/or Bmem to COVID-19, and thus provide a premise for continued vaccination and the use of measures of both arms of the immune system to assess vaccine efficacy.

## METHODS

### Participants

In 2021, 29 healthy adults and 25 patients with IEI were recruited in a low-risk observational research study, after they had chosen vaccination against COVID-19 (**Table 1, Supplementary Tables 1, 2 &3**). Blood samples were obtained at 1-month post doses 2 and 3.^14–16^ In addition, basic demographics including age, sex, clinical diagnosis, COVID-19 infection status, COVID-19 vaccination formulations, and treatment status were collected throughout the study. The 29 healthy adults were selected from the previously-reported cohort, of which sufficient residual stored, live cells remained.^14–16^ Exclusion criteria for healthy adults were the presence of hematological or immunological disease or receiving systemic immunosuppressive therapies. The study was conducted according to the Declaration of Helsinki and approved by local ethics committees (Alfred Health ethics no. 32/21, Monash University project no. 72794).

**Table 1:** Characteristics of IEI patients and healthy controls.

|  | **Patients with IEI**<br>**(n=25)** | **Controls**<br>**(n=29)** | **P Value** |
| --- | --- | --- | --- |
| **DEMOGRAPHICS** |  |  |  |
| Sex: female n (%) | 72% (18/25) | 72% (21/29) | >0.99<sup>1</sup> |
| Age (y), median (range) | 48 (23-66) | 38 (24-62) | 0.33<sup>2</sup> |
| **IEI diagnosis, n (%)** |  |  |  |
| CD4 T cell cytopenia | 4% (1/25) | - | - |
| CID | 12% (3/25) | - | - |
| PAD | 76% (19/25) | - | - |
| PU.MA | 4% (1/25) | - | - |
| CVID | 40% (10/25) | - | - |
| HGG | 8% (2/25) | - | - |
| SpAD | 16% (4/25) | - | - |
| NFKB1-haplo | 8% (2/25) | - | - |
| XLP2 | 4% (1/25) | - | - |
| CHAI | 4% (1/25) | - | - |
| **IMMUNOLOGICAL PRESENTATION** |  |  |  |
| Decreased serum immunoglobulin levels* |  |  |  |
| IgG | 75% (18/24) | N/A | - |
| IgA | 56% (14/25) | N/A | - |
| IgM | 44% (11/25) | N/A | - |
| Reduced cell numbers<sup>#</sup> |  |  |  |
| B cells | 12% (3/25) | N/A | - |
| CD4<sup>+</sup> T cells | 28% (7/25) | N/A | - |
| CD8<sup>+</sup> T cells | 44% (11/25) | N/A | - |
| **TREATMENT** |  |  |  |
| IgRT, n (%) | 76% (19/25) | - | - |
| Immunomodulatory medications | 52% (13/25) | - | - |
| COVID-19 infection <sup>δ</sup> | 8% (2/25) | 0% (0/29) | 0.12 |
| **VACCINATION CHARACTERISTICS** |  |  |  |
| Vaccine formulation for doses 1 and 2 |  |  | 0.08<sup>1</sup> |
| BNT162b2 (mRNA) | 72% (18/25) | 48% (14/29) |  |
| ChAdOx1 (Adenoviral) | 28% (7/25) | 52% (15/29) |  |
| Vaccine formulation for dose 3 |  |  | 0.23<sup>1</sup> |
| BNT162b2 | 88% (22/25) | 97% (28/29) |  |
| mRNA-1273 | 12% (3/25) | 3% (1/29) |  |

\*before IgRT and defined as below reference range.<sup>55</sup> <sup>#</sup> defined as below 5<sup>th</sup> percentile of controls.<sup>55</sup>
<sup>δ</sup> All recorded between 1m post-dose 2 bleed and before receiving vaccination dose 3. CHAI, Cytotoxic T lymphocyte antigen 4 haploinsufficiency with autoimmune infiltration; CID, combined humoral and cellular immunodeficiency; CVID, common variable immunodeficiency; HGG, hypogammaglobulinemia; IgRT, immunoglobulin replacement therapy; NFKB1, nuclear factor kappa-B subunit 1; NIC, non-infectious complications; PAD, predominantly antibody deficiency; PU.MA, PU.1-mutated agammaglobulinemia; SpAD, specific antibody deficiency; XLP2, X-linked lymphoproliferative syndrome 2. <sup>1</sup> Chi-squared test. <sup>2</sup> Mann-Whitney test.

### Measurement of neutralizing titers of SARS-CoV-2 antibodies in plasma

The live virus neutralization assay was performed, as previously described.^18, 52^ Clonal HAT-24 cells were generated by transducing lentiviral particles into HEK293T (Thermo Fisher, R70007) cells to stably express ACE2 and TMPRSS2 receptors. The cells were trypsinized, resuspended in DMEM-5% foetal bovine serum (FBS) and stained with Hoechst-33342 (5% v/v) live nuclear dye (Invitrogen, R37605) before seeding in a 384-well plate at a density of 1.6 × 10^4^ cells per well. Test samples were serially diluted (2-fold) with DMEM-5% FBS and an equal volume of SARS-CoV-2 virus solution, at twice the median lethal dose, was added to the diluted sample. Sample-virus mixture was incubated for 1hr at 37°C and 40 μl per well was then added to the plated cells. Plates were incubated for 20 hours at 37°C, 5% CO2 before enumerating nuclear counts with high-content fluorescence microscopy using Cytiva InCell Analyzer HS2500 and IN Carta software. Percentage of virus neutralization was calculated with the formula %N = (D-(1-Q)) x 100/D, where Q=nuclei count / average count for uninfected controls, and D=1-Q for average count for infected controls). Sigmoidal dose-response curves and interpolated IC50 values (reciprocal dilution at which 50% neutralization is achieved) were determined using Sigmoidal, 4PL model of regression analysis in GraphPad Prism (GraphPad Software, USA). The lowest amount of neutralizing antibody detectable is a titer of 20. All samples that did not reach 50% neutralization were assigned an arbitrary value of 10.

### Protein production and tetramerization

SARS-CoV-2 ancestral Wuhan-Hu-1 (WH1) spike RBD and nucleocapsid (NCP) proteins, as well as Omicron BA.2 and BA.5 subvariant RBDs were recombinantly produced with an N-terminal leader sequence, and a C-terminal AviTag for biotin ligase (BirA)-catalyzed biotinylation, and 6-His affinity tag for cobalt affinity column purification, as described previously.^14–16, 53^ All proteins were purified and used for ELISAs. Aliquots of the purified RBD proteins were biotinylated and tetramerized with fluorochrome-conjugated streptavidins for flow cytometry at a protein:streptavidin molar ratio of 4:1 to form: [RBD WH1]_4_-BUV395, [RBD WH1]_4_-BV421, [RBD BA.2]_4_-BUV737 and [RBD BA.5]_4_-BV480 (**Supplementary Tables 4, 5 and 6**).

### ELISA

ELISAs were performed for quantitation of plasma IgG specific for ancestral NCP and ancestral, Omicron BA.2 and BA.5 RBDs. 96-well EIA-RIA plates (Corning Incorporated, Costar, St Louis, MO) were coated with 2 μg/ml recombinant unbiotinylated protein monomers overnight at 4°C. Separate wells were serially diluted with recombinant human IgG (in-house made human Rituximab), to generate a standard curve for IgG level quantitation. Plates were then blocked with 3% BSA in PBS and subsequently incubated with plasma samples diluted 1:30 to 1:10,000 for quantitation of RBD-specific IgG, and 1:300 for NCP-specific IgG post-dose 2 and post-dose 3 of COVID-19 vaccination. No plasma was added to standard curve wells or His control wells. Antigen-specific IgG was detected using rabbit anti-human IgG HRP (Dako, Glostrup, Denmark). ELISA plates were developed using TMB solution (Life Technologies, Carlsbad, CA), and the reaction was stopped with 1M HCl. Absorbance (OD450nm) was measured using a Multiskan Microplate Spectrophotometer (Thermo Fisher Scientific). Titration curves were generated, and the area under the curve (AUC) was calculated for each variant calculated using GraphPad Prism 10. IgG binding to Omicron variant RBD was quantified relative to ancestral WH1 as a percentage of the WH1 AUC.^14–16, 53, 54^

### Sample processing and Flow cytometry

#### Assessment of absolute numbers of leukocyte and lymphocyte subsets

Blood samples were processed, as described previously.^14–16, 22, 53–55^ Absolute numbers of major leukocyte subsets were determined, as previously described, using a lyse-no-wash method within 24hrs of blood sampling in Vacutainers containing EDTA (BD Biosciences, San Jose, CA, USA).^14–16, 22, 53–55^ Briefly, 50 μl whole blood was added to a BD Trucount tube (BD Biosciences) and incubated with an antibody cocktail of 20µl to stain CD3, CD4, CD8, CD16, CD45 and CD56 for 15 min in the dark at room temperature (**Supplementary Tables 4 and 6**). Subsequently, cells were incubated with 1X BD Lysis Solution (BD Biosciences) for 15 min to lyze blood cells. Samples were acquired on the BD FACSLyric analyzer, and data were analyzed using FlowJoTM Software v.10.8.1 (BD Biosciences). Trucount data were used to calculate absolute numbers of RBD-specific Bmem,^14–16, 22, 53, 54^ and spike-specific Tmem subsets. The remainder of the sample was used to separate and store plasma (-80°C), and to isolate live peripheral blood mononuclear cells (PBMC) following Ficoll-paque density gradient centrifugation and cryopreservation in liquid nitrogen for later analysis of spike-specific T cells and RBD-specific B cells.

#### Immunophenotyping of RBD-specific memory B cells

For the detection of antigen-specific Bmem, cryopreserved PBMC were thawed and stained, as previously described.^14–16, 22, 53, 54^ Briefly, 10-15 million PBMC were incubated with fixable viability stain 700 (BD Biosciences), antibodies against CD3, CD19, CD21, CD27, CD38, CD71, IgA, IgD, IgG1, IgG2, IgG3, IgG4 (**Supplementary Tables 5 and 6**), and 5 μg/ml each of [RBD WH1]4-BUV395, [RBD WH1]4-BV421, and [RBD BA.2]4-BUV737 and [RBD BA.5]4-BV480 for 15 minutes at room temperature in a total volume of 250μl FACS buffer (0.1% sodium azide, 0.2% BSA in PBS). In parallel, 5 million PBMC were incubated with fixable viability stain 700 (BD Biosciences), antibodies against CD3, CD19, CD27 and IgD, and BUV395-, BUV737-, BV421- and BV480-conjugated streptavidin controls (**Supplementary Table 5 and 6**). Following staining, cells were washed with FACS buffer, fixed with 2% paraformaldehyde for 20 minutes at room temperature and washed once more. Following filtration through a 70 μM filter, cells were acquired on a 5-laser LSRFortessa X-20 (BD Biosciences) or 4L Cytek Aurora. Flow cytometer set-up and calibration was performed using standardized EuroFlow SOPs, as previously described.^14–16, 22, 53, 54^ Data analysis was performed using FlowJo^TM^ Software v.10.8.1 (BD Biosciences).

#### Immunophenotyping of Spike-specific T cells

Spike-specific T cells were enumerated and characterized based on a previously reported protocol.^56^ Thawed PBMCs were seeded at 1.5 x 10^6^ cells per well and cultured under three different conditions in a 96-well flat-bottom plate: 1) 1μg/ml plate-bound CD3 (clone OKT3) and 1μg/ml soluble CD28 (clone CD28.2) monoclonal antibodies (mAb eBioscience); 2) final concentration 0.6nM/mL PepTivator® peptide (SARS-CoV-2 Protein_S Complete, research grade (Miltenyi Biotec), and 3) Media alone (RPMI supplemented with 10% FCS and 1% Pen/Strep, 2-Mercaptoenthanol). CD3 mAb was immobilized by coating wells for 2 hours at 37°C and washed away before addition of cells. Assay plates were incubated for 4 hours at 37°C, and then for a further 16 hours in the presence of Brefeldin A (3 µg/ml, Golgi Plug®). After stimulation, the cells were washed in PBS and subsequently resuspended in brilliant stain buffer and Fc receptor block (BD Biosciences) which was diluted in FACS buffer (PBS with 0.1% sodium azide and 0.2% BSA). Three wells were used for peptide-specific stimulation and pooled subsequently to acquire sufficient numbers of peptide-specific T cells. To minimize background staining, the samples were briefly incubated for 10 minutes at RT.

The cell viability dye (FVS780) was added in PBS, followed by an FACs buffer containing an antibody cocktail of CD3 HorV500, CD4 BV786, CD8α R718, CCR7 BV605, CD45RA BV711 and CXCR5 BV421 (BD Biosciences) (**Supplementary Tables 3 and 6**). Samples were incubated for 20 minutes in the dark at RT, washed once in FACs buffer and centrifuged in 100µl of brilliant stain buffer (BD). Supernatants were aspirated and cell pellets resuspended in 200µl of BD cytofix/cytoperm (BD Biosciences) to permeabilize cell membranes. Samples were incubated for a further 20 minutes at room temperature and then centrifuged. Cell pellets were resuspended in 200µl of Phosphoflow Perm/Wash Buffer I diluted 10x (BD Biosciences) in milliQ water, centrifuged and resuspended with the intracellular antibody mix containing CD134 PE, CD137 APC, CD69 PE-Cy7, IFNγ FITC and TNFα PerCP Cy5.5 in phosphoflow perm/wash buffer I (BD Biosciences (**Supplementary Tables 3 and 6**). After a 20-minute incubation at RT in the dark, samples were washed twice in the presence of Phosphoflow Perm/Wash Buffer I. Final cell pellets were resuspended in chilled PBS and filtered through a 70 µM cell strainer (BD Biosciences) into FACS tubes. All samples were stored overnight at 4°C and then acquired on a BD FACSLyric™ analyzer using pre-determined “T-cell assay” reference settings. Cytometer performance and settings were monitored before each run using BD CS&T RUO beads (BD Biosciences). Up to 100,000 events were acquired for positive and negative control samples, and 1-3 million events for peptide-specific samples.

### Data analysis and statistics

All flow cytometry data were analyzed with FlowJo v10 software (BD Biosciences). Statistical analysis was performed with GraphPad Prism 10 Software (GraphPad Software). Matched pairs were analyzed with the non-parametric Wilcoxon matched pairs signed rank test with Bonferroni correction for multiple comparisons. Comparisons between 3 or more groups were performed using the Friedman’s test (paired) or Kruskal-Wallis (unpaired) with Dunn multiple comparisons test. Statistical analysis for qualitative data was performed with the Chi-squared test. For all tests, *p* < 0.05 was considered significant.

## RESULTS

### Cohort Characteristics

29 healthy adult healthcare workers (median age 38, range 24-62 years; 72% female) and 25 IEI patients (median age 48, range 23-66 years; 72% female) were included in the study (**Table 1 and Supplementary Table 1& 2**). All study volunteers received either three doses of mRNA vaccine (homologous; n=14 healthy controls and n=18 IEI patients), or two doses of an adenoviral ChAdOx1 vaccine, followed by a third dose mRNA booster (heterologous; n=15 healthy controls and n=7 IEI patients). Of the 54 third doses, one control and three patients received monovalent mRNA-1273, whilst the remaining 50 received monovalent BNT162b2 (**Table 1**). All controls evaluated have been reported previously.^14–16^ There were no significant differences in the age or sex distribution between groups, except for the heterologous IEI group, who were a median of 20 years older than controls (median age of 58 years vs 38 years, p= 0.0003) (**Supplementary Table 2**).

Peripheral blood was collected at 4 weeks after both dose 2 (D2; controls 28 (25-35) days; IEI patients 30 (20-148) days) and dose 3 (D3; controls 29 (27-43) days; IEI patients 30 (25-169) days). There was no significant difference in the interval between doses 1 and 2 (**Fig 1 A**), or the time from vaccination to sample collection at each time point regardless of vaccine regimen received (**Table 1** and **Supplementary Table 2**). However, due to presumed susceptibility of severe SARS-CoV-2 infection,^6^ Australian policy changed after 2021 to include a 3-dose primary vaccine regimen, which is reflected in the significantly shorter interval between doses 2 and 3 in IEI patients (median, 112 days) compared to controls (median, 184 days) (**Fig 1 A** and **Supplementary Table 2**).^11^

**FIG 1.**
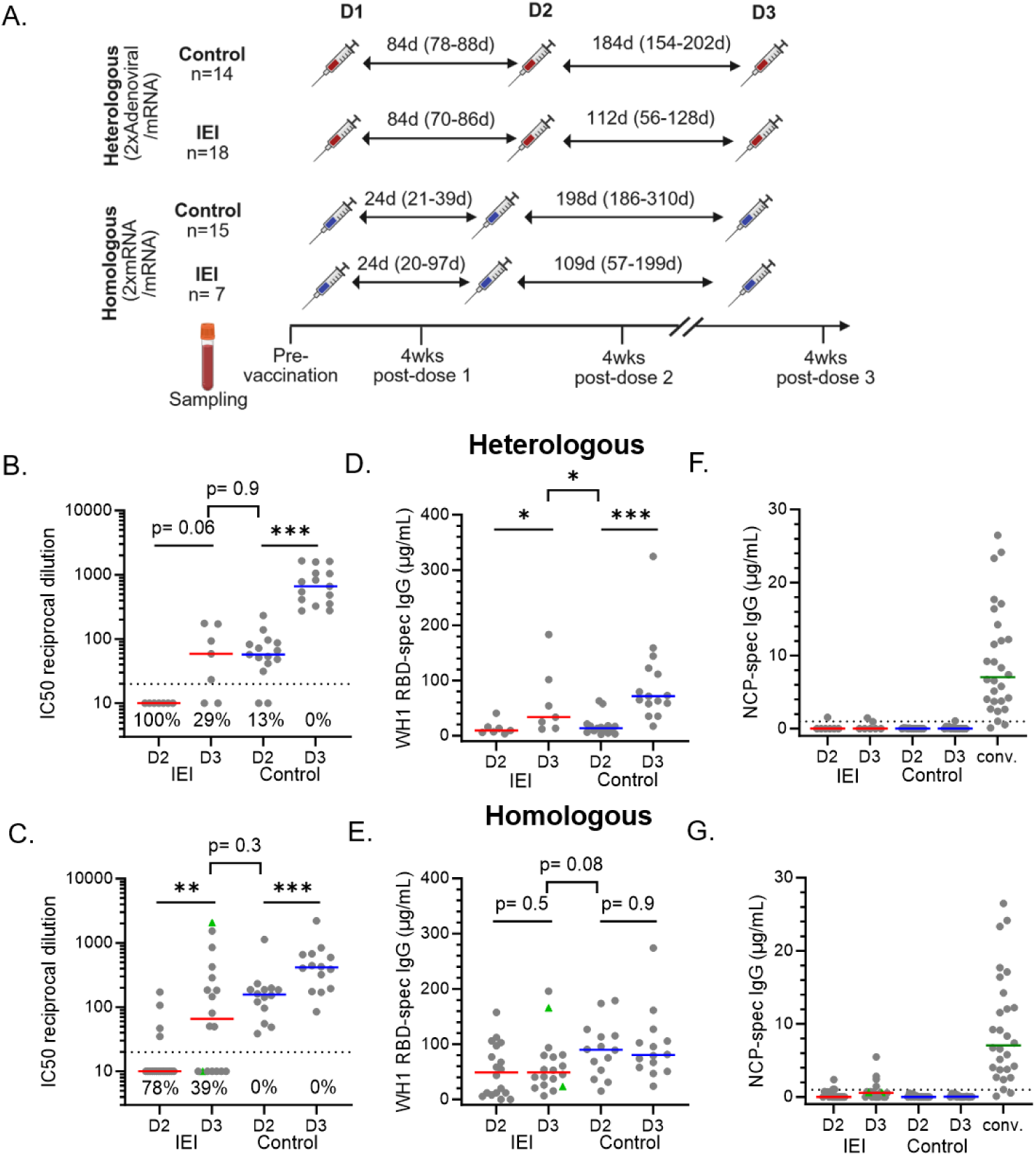
Ancestral SARS-CoV-2-specific antibody responses after 2 and 3 COVID-19 vaccine doses in patients with IEI. (**A**) Schematics of patient cohorts, vaccination schedules and sampling timepoints (**Supplementary Tables 1 & 2**). Samples included 1-month post-dose 2 and 1-month post-dose 3 samples from IEI patients and healthy controls receiving primary ChAdOx1 (heterologous) or BNT162b2 (homologous) vaccination followed by a 3^rd^ dose mRNA vaccine. NAb titers to (**B**) heterologous and (**C**) homologous vaccination. The dotted lane indicates the neutralization cut-off (IC50 = 20).^22^ WH1 RBD-specific for (**D**) heterologous and (**E**) homologous vaccine recipients, and WH1 NCP-specific IgG response in (**F**) heterologous and (**G**) homologous recipients, including convalescent individuals(6-120 days post COVID-19 infection).^22^ Green triangles denote confirmed breakthrough infections (BTI) prior to sampling (**Supplementary Tables 3**). Red and blue lines represent median values. Mann-Whitney test and Wilcoxon matched-pairs signed rank test used for unpaired and paired data, respectively (*p<0.05, **p<0.001, ***p<0.0001). Healthy donor data previously published in ^14–16^.

The majority of IEI patients (n=19; 76%) were classified as having a predominantly antibody deficiency (PAD; category 3), and included diagnoses of PU.1-mutated agammaglobulinemia (PU.MA) due to a deletion in the *SPI1* gene (4%),^57^ CVID (40%), hypogammaglobulinemia (8%), SpAD (16%). Furthermore, 8% of our patients had immune dysregulation (category 4), 12% CID (category 1) and 4% an uncategorized IEI (CD4 T-cell cytopenia).^2, 7, 58^ Of the 25 patients included in this study, 15 were sequenced with 11 receiving no result, and the remaining four patients (16%) returning a genetic diagnosis: CTLA-4 haploinsufficiency with autoimmune infiltration (CHAI; n=1)^59^, *NFKB1* haploinsufficiency (n=2) and X-linked lymphoproliferative syndrome-2 (XLP-2) due to a variant in *XIAP* (n=1)^60^ (**Table 1 and Supplementary Table 1**).

Most patients have a history of infectious disease (92%) (**Table 2** and **Supplementary Table 3**), and these typically involved the respiratory tract: sinusitis (80%), pneumonia (16%), otitis media (12%), bronchiectasis (16%), as well as the gastrointestinal tract (8%) and other sites (52%). Furthermore, 76% of patients presented with at least one non-infectious complication, including autoimmunity (40%), gastrointestinal disease (28%), asthma (28%) and atopy/allergy (16%) (**Table 2** and **Supplementary Table 3**). At the time of inclusion in the study, 19 patients (76%) received IgRT, and 13 patients (52%) were prescribed immunomodulators for treatment of asthma, autoimmunity or IgRT tolerability (**Table 1 and Supplementary Table 1**).

**Table 2:**
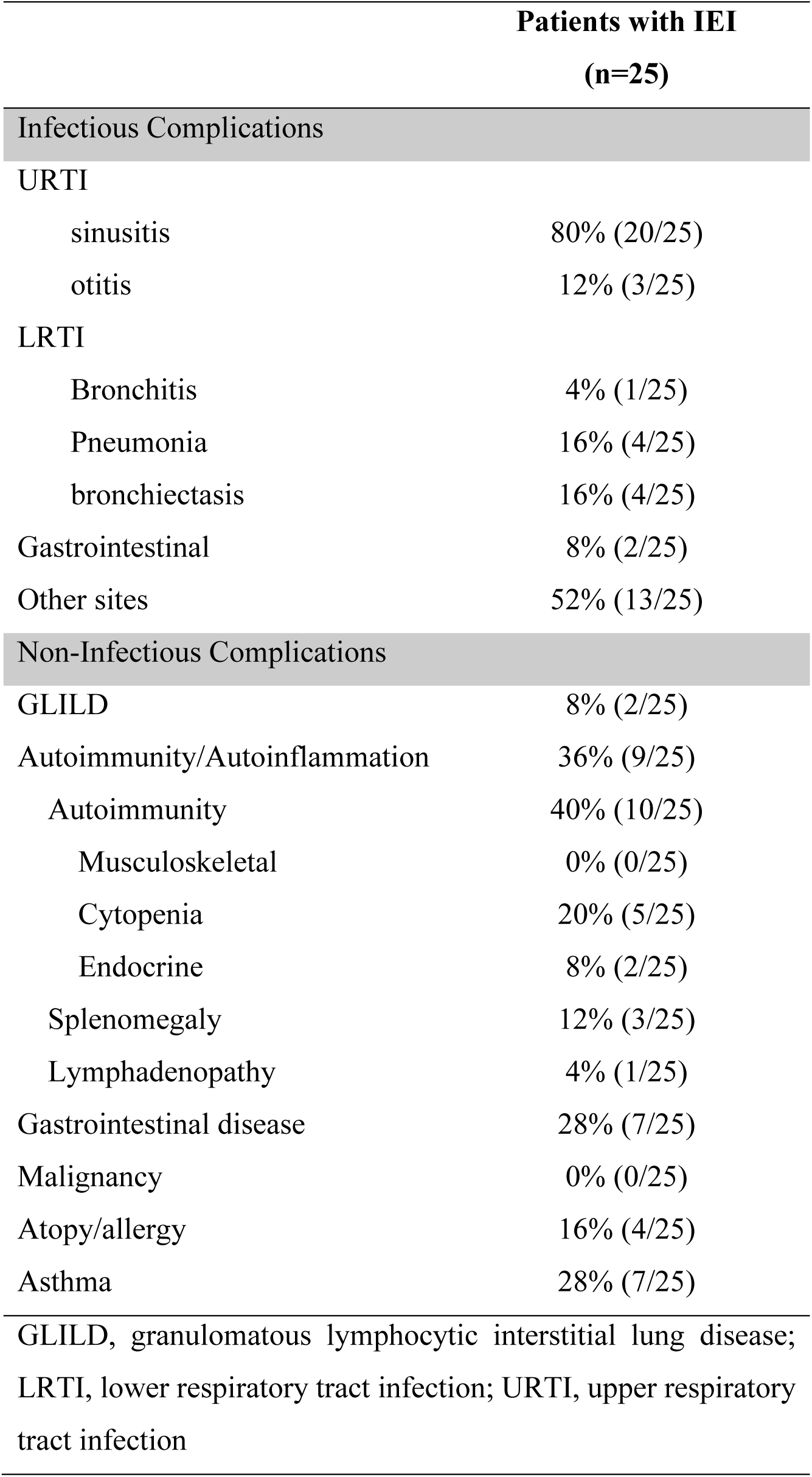
Comorbidities of patients with Inborn Errors of Immunity.

Serum IgG levels at diagnosis were available from 24 patients, and were reduced in 18 (75%). Serum IgA was reduced in 14/25 patients (56%) and IgM was reduced in 11/25 (44%) patients (**Table 1 and Supplementary Table 1**). ^55^

### Reduced antibody response in IEI patients to the third COVID-19 vaccine dose

To evaluate the antibody responses to vaccination, NAb to Ancestral SARS-CoV-2 virus were determined 1-month after doses 2 and 3. As antibody responses following adenoviral vaccination are reported to be 8-10 times lower than to mRNA vaccination,^14, 15^ the control and IEI cohorts were stratified based on primary vaccine formulation. After dose 2, 13/15 (87%) healthy controls had detectable NAb titres, which significantly increased to 15/15 (100%) after dose 3 (**Fig 1 B&C**). In contrast, two vaccine doses only elicited NAb in 4/25 (16%) IEI patients, which significantly increased after dose 3, yet a significant proportion failed to form NAb after dose 3 (29% heterologous versus 39% homologous) (**Fig 1 B&C**).

In parallel, IgG binding levels to WH1 Spike RBD were significantly lower in IEI patients than in controls after both vaccine doses (**Fig 1D&E**). As shown previously for healthy controls, ^15^ following dose 3, WH1 RBD-specific IgG levels were only significantly increased in the heterologous vaccine groups (7/7 IEI patients and 14/15 controls), raising these to similar levels as observed in the homologous vaccine groups (**Fig 1 D**).

The presence of SARS-CoV-2 NCP-specific IgG can be indicative of a previous breakthrough infection (BTI).^14, 16, 22^ No BTIs were observed in our healthy control group,^15^ and 2/25 IEI patients self-reported a SARS-CoV-2 BTI in-between doses 2 and 3 of vaccination presenting a clinically mild infection (**Table 1** and **Supplementary Tables 1**). This was confirmed by positive NCP IgG serology in the post-dose 3 samples (**Fig 1 F&G**).

### Impaired Induction of Spike Specific CD8^+^ T cells in IEI patients

Consistent with diagnostic evaluations (**Table 1**), total CD4^+^ and CD8^+^ T-cell numbers were significantly lower in IEI patients than in controls, with 28% of patients showing reduced CD4^+^ T-cell numbers and 44% with reduced CD8^+^ T-cell numbers (**Supplementary Fig 1**). This was due to significantly lower naive CD4^+^ and CD8^+^ T-cell, CD4^+^ central memory (T_CM_) and CD4^+^ effector memory CD45RA revertant T-cell (T_EMRA_) counts (**Supplementary Fig 1**).

To evaluate SARS-CoV-2-specific T cells, PBMC were stimulated *in vitro* with overlapping peptides of the SARS-CoV-2 Spike protein, and analyzed by flowcytometry for expression of Antigen-Induced Markers (AIM) CD69, CD134 (OX-40) and CD137 (4-1BB) (**Supplementary Fig 2 A-C**). The threshold for gating of positive events was set on the basis of the unstimulated controls, which consistently contained cells with dim expression of CD137. Following stimulation, CD8^+^ T cells expressing CD69 and CD137 were clearly defined over background (unstimulated cells) (**Fig 2 A & Supplementary Fig 2 A**). Spike-specific CD8^+^ memory cells were detectable at similar numbers in both groups, with no significant differences between doses (**Fig 2 B**) or vaccine regimens (**Supplementary Fig 3 A&B**). There was a large interindividual variation with some patients showing 10-20-fold higher numbers than the cohort median (**Fig 2 B**).

**FIG 2.**
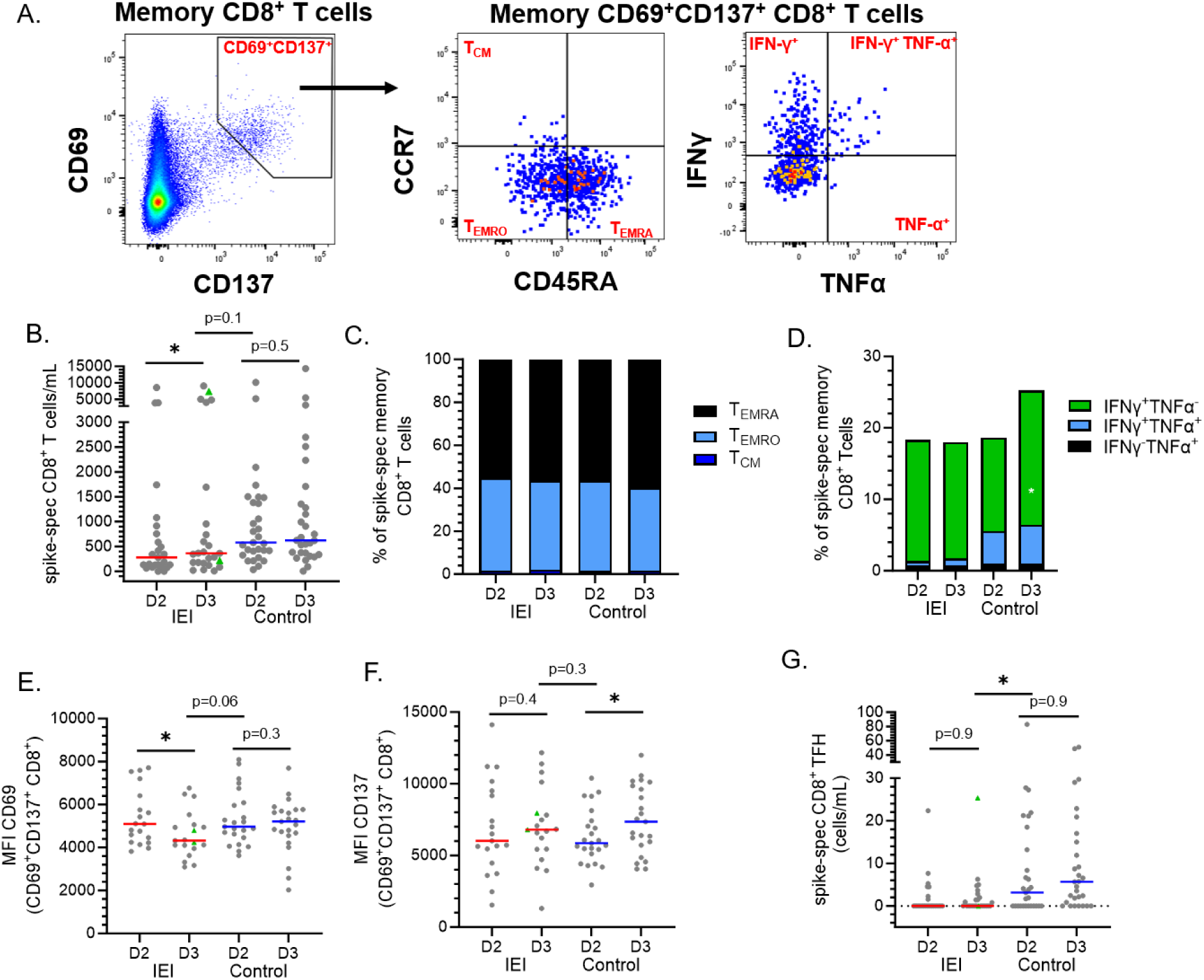
Spike-specific CD8^+^ T cells in IEI patients after 2 or 3 COVID-19 vaccine doses. (**A**) Gating strategy for spike-specific CD8^+^ Tmem and cytokine expression therein. (**B**) Spike-specific CD8^+^ Tmem numbers after vaccine doses 2 and 3 in IEI patients and controls. (**C**) Distribution of spike-specific CD8^+^ Tmem subsets. (**D**) Frequencies of Spike-specific CD8^+^ T cells with intracellular expression of IFN-γ and/or TNF-α. (**E**) CD69 and (**F**) CD137 expression levels on Spike-specific CD8^+^ Tmem. (**H**) Spike-specific CD8^+^ Tfh numbers. Green triangles indicate breakthrough infection (BTI) before sampling (**Supplementary Table 3**). Red and blue lines represent median values. Mann-Whitney test for unpaired data and Wilcoxon matched-pairs signed rank test for paired data. *p<0.05, **p<0.001, ***p<0.0001.

To further examine the phenotype of CD69^+^CD137^+^ CD8^+^ T cells, we determined the distribution of Spike-specific CD8^+^ T cells within the effector and central memory compartments, and intracellular IFN-γ and TNF-α expression (**Fig 2 A**). Regardless of time point, almost 60% of Spike-specific CD8^+^ T cells were T_EMRA_ in both cohorts, with the remainder being mostly T_EMRO_ and <5% showing a T_CM_ phenotype. (**Fig 2 C**). Of the Spike-specific CD8^+^ T cells, 18-25% expressed IFN-γ and/or TNF-α in both cohorts. Of note, TNF-α ^+^ proportions are significantly lower in IEI patients after both doses, with IFN-γ^+^ proportions significantly higher in controls after dose 3 (**Fig 2 D**). Whilst AIM markers clearly distinguished activated T-cell populations in both cohorts, it was evident that CD69 expression was lower in patients after dose 3 (**Fig 2 E**) suggesting a reduced activation capacity. This was not observed when examining CD137 expression (**Fig 2 F**).

CD8^+^ Tfh cells reside within B cell follicles and are thought to promote B-cell antibody class-switching alongside CD4^+^ Tfh cells.^61, 62^ Numbers of circulating Spike-specific CD8^+^ Tfh were significantly higher in controls than IEI patients at both timepoints (**Fig 2 G**). In fact, most patients (19/25, 76%) failed to develop detectable spike-specific CD8^+^ T_FH_ cells, compared to 41% of healthy controls. A third vaccine dose increased detection rates in both groups to 44% in IEI patients and 76% in controls (**Fig 2 G**).

Overall, these findings suggest that, upon COVID-19 vaccination, Spike-specific CD8^+^ Tmem are formed. However, in IEI patients these displayed impairments in their activation profile.

### Altered formation of vaccine-specific CD4^+^ T cells

In parallel to CD8, we evaluated Spike-specific memory CD4^+^ T cells following stimulation with overlapping peptides of the Spike protein (**Fig 3 A &** (**Supplementary Fig 2 B & C**). Unlike the homogeneous expression of CD137 and CD69, the expression of CD134 was notably heterogeneous in Spike-specific CD4^+^ T cells (**Supplementary Fig 2C**). Given peak expression of CD134 is not observed until 48 hours post-stimulation,^56^ we defined our AIM CD4^+^ T cells based on co-expression of CD137 and CD69. Numbers of SARS-CoV-2 Spike specific CD4^+^ T cells were similar in both cohorts after doses 2 and 3 (**Fig 3 A&B, Supplementary Fig 2 D**) and regardless of vaccine regimen (**Supplementary Fig 3 C&D**). Notably, several patients displayed higher numbers of Spike-specific CD4^+^ T cells, showing the high heterogeneity in the cohort (**Fig 3 B**). The majority of SARS-CoV-2 Spike-specific CD4^+^ T cells displayed a T_EMRO_ phenotype in both groups at both timepoints (**Fig 3 C**). The T_EMRO_ subset significantly expanded after dose 3 at the expense of T_CM_ in controls, whereas a similar trend was not observed in IEI patients (**Fig 3 C**).

**FIG 3.**
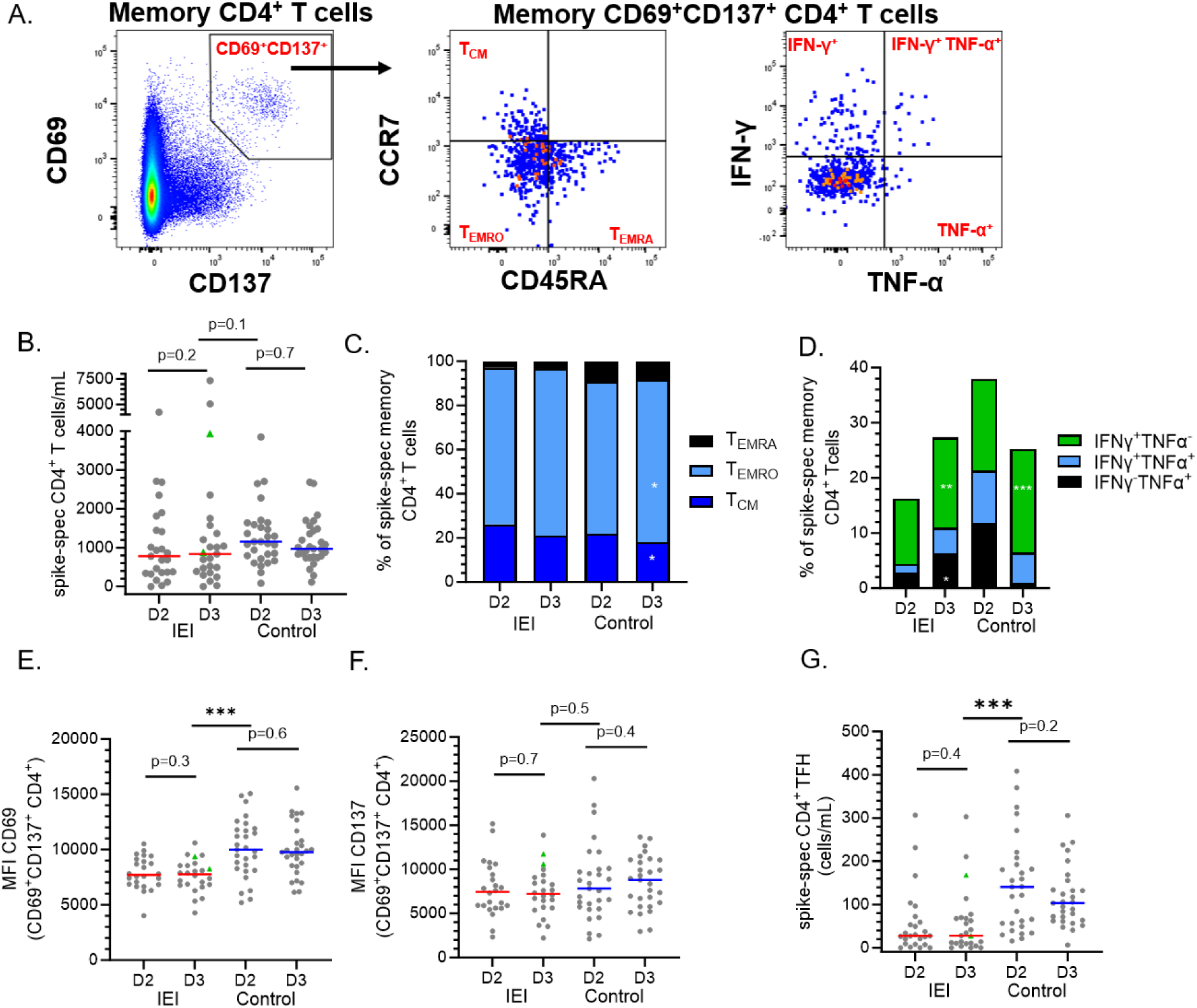
Spike-specific CD4^+^ T cells after 2 or 3 COVID-19 vaccine doses. (**A**) Gating strategy for spike-specific CD4^+^ T Tmem and cytokine expression therein. (**B**) **S**pike-specific CD4^+^ Tmem numbers after vaccine doses 2 and 3 in IEI patients and controls. (**C**) Distribution of spike-specific CD4^+^ Tmem subsets. (**D**) Frequencies of Spike-specific CD4^+^ T cells with intracellular expression of IFN-γ and/or TNF-α. (**E**) CD69 and (**F**) CD137 expression levels on Spike-specific CD4^+^ Tmem. (**H**) Spike-specific CD4^+^ Tfh cell numbers. Green triangles indicate breakthrough infection (BTI) before sampling (**Supplementary Table 3**). Red and blue lines represent median values. Mann-Whitney test used for unpaired data and Wilcoxon matched-pairs signed rank test for paired data. *p<0.05, **p<0.001, ***p<0.0001.

We assessed the functional capabilities of Spike-specific CD4^+^ T cells by examining the intracellular cytokine profiles (**Fig 3 D**). Following dose 2, in IEI patients about 16% of Spike-specific CD4^+^ T cells expressed IFN-γ and/or TNF-α, and this was increased to 27% after dose 3. In contrast, 37% of Spike-specific CD4^+^ T cells of healthy controls expressed IFN-γ and/or TNF-α, and this was increased to 25% after dose 3 (**Fig 3 D**). IFN-γ^+^TNF-α^+^ are significantly higher in controls than patients after dose 2, whereas IFN-γ^+^ are significantly higher after dose 3. As seen for CD8^+^ T cells, the median fluorescence intensities of CD69 were significantly lower than controls after both vaccine doses (**Fig E**), whereas no differences were observed for CD137 or CD134 (**Fig 3 F & Supplementary Fig 2 E&F**).

Total CD4^+^ Tfh cell numbers, defined as CXCR5^+^CD45RA^−^, were not different between patients and controls. In contrast, we observed that IEI patients had 7-fold fewer Spike-specific Tfh cells than healthy controls after both doses (**Fig 3 G**). After dose 3 Tfh cell numbers were unchanged in the patient cohort (**Fig 3 G**), and while absolute numbers of Spike-specific Tfh cells dropped marginally in the controls post-dose 3, these were still 5-fold higher than in patients at the equivalent dose. Overall, these findings suggest that IEI patients mount CD4^+^ T-cell memory after doses 2 and 3 of COVID-19 vaccination. However, Spike-specific CD4^+^ T cells in the IEI patients displayed quantitative impairments in activation, cytokine production and Tfh formation

### Reduced formation of Spike RBD-specific memory B cells in patients with IEI

WH1 RBD-specific Bmem were evaluated within the total CD19^+^ Bmem population after exclusion of CD27^-^IgD^+^ naive B cells (**Fig 4 A and Supplementary Fig 4**). Prior to Bmem analysis, we identified two patients lacking circulating B cells and/or Bmem, who were excluded from further analysis (**Supplementary Fig 5 A**). Consistent with diagnostic evaluation (**Table 1**), total B-cell numbers were reduced in 16% of IEI patients after both doses (**Supplementary Fig 5 B&C**).

**FIG 4.**
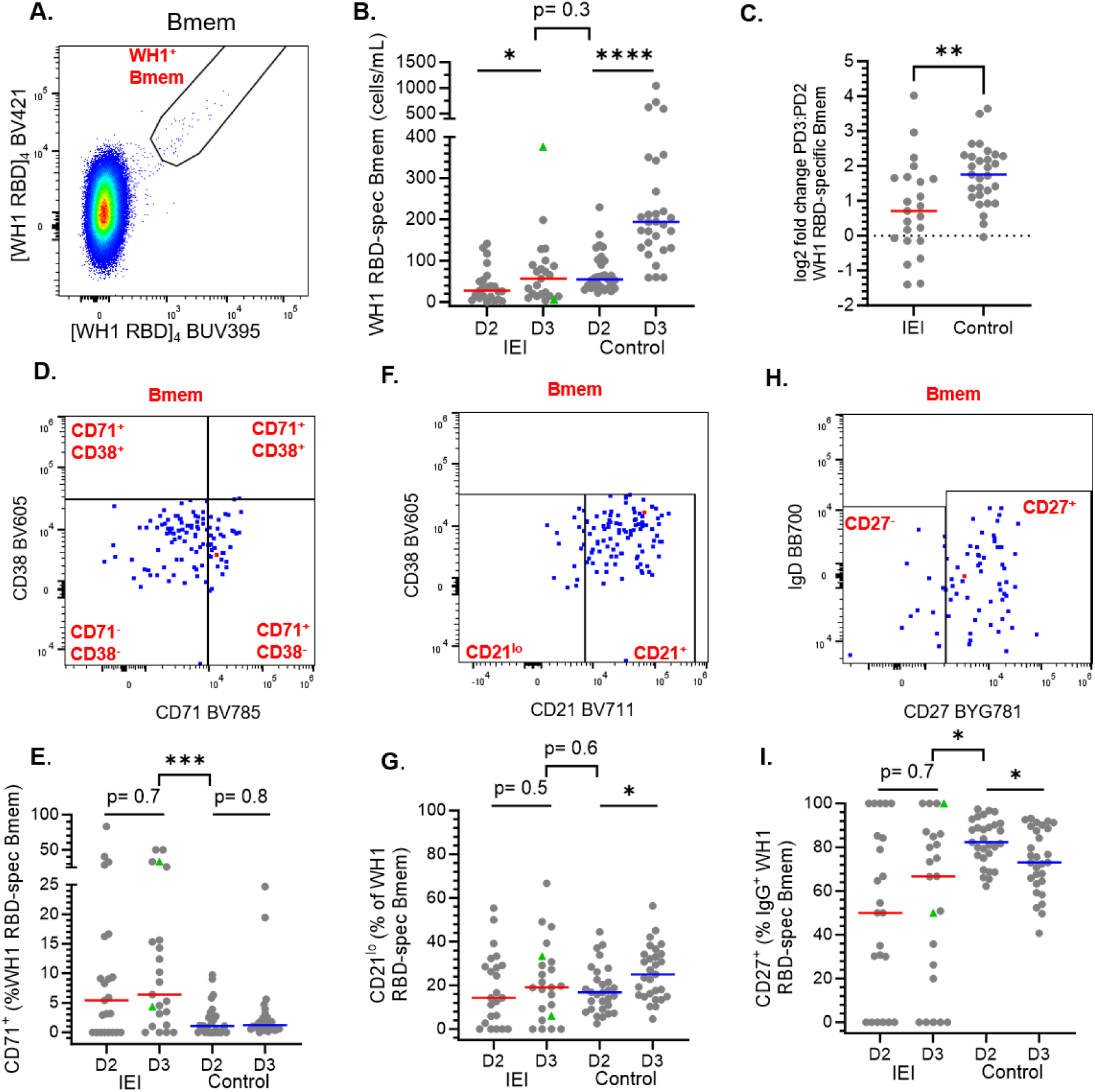
Formation and maturation of ancestral RBD-specific Bmem after 2 and 3 COVID-19 vaccine doses in IEI patients. (**A**) Gating strategy of antigen-specific B cells within Bmem from a representative IEI patient. (**B**) WH1 RBD-specific Bmem numbers in IEI patients and controls after vaccine doses 2 and 3. (**C**) Fold change of WH1 RBD-specific Bmem numbers between D3 and D2. (**D, E**) CD38^dim^CD71^+^ activated cells within RBD-specific Bmem. (**F, G**) CD21^lo^CD38^dim^ cells within RBD-specific Bmem. (**H, I**) CD27^+^ cells within IgG^+^ RBD-specific Bmem. Green triangles represent individuals with confirmed breakthrough infection (BTI) before sampling (**Supplementary Table 3**). Red and blue lines represent median values. Mann-Whitney test was performed for unpaired data and Wilcoxon matched-pairs signed rank test for paired data. *p<0.05, **p<0.001, ***p<0.0001. Healthy donor data previously published in ^14–16^.

WH1 RBD-specific Bmem were readily identified in IEI patients using recombinant protein tetramers,^14–16, 22, 54^ although the numbers were significantly lower than in healthy controls after both doses (**Fig 4 B**) with no significant differences observed between regimens (**Supplementary Fig 3 E&F**). Following dose 3, the numbers of WH1 RBD-specific Bmem significantly increased in both patients and controls (**Fig 4 B**). However, boosting was less in IEI patients (2.7-fold) than in controls (4-fold) (**Fig 4 C**). This is a direct result of dose 3 boosting numbers of WH1 RBD-specific Bmem in 70% IEI patients compared to 97% of controls (**Fig 4 B&C**). Of note, one of the patients with a confirmed BTI exhibited the highest WH1 RBD-specific Bmem numbers (**Fig 4 B**).

### Expansion of CD27^+^ IgG^+^ WH1 RBD-specific Bmem in IEI patients 1-month after dose 3

To define fractions of WH1 RBD-specific Bmem that show signs of recent activation, as in previous studies we evaluated the cell surface expression of CD71, CD21 and CD27 (**Fig 4 D-H**).^14–16, 22, 54^ After both doses 2 and 3, higher frequencies of WH1 RBD-specific Bmem expressed CD71 in IEI patients than in controls (**Fig 4 D&E**). This was not associated with evidence of a BTI (**Fig 4 E**), or differences in total Bmem expressing CD71 in IEI patients (**Supplementary Fig 5 D&E**), and could indicate sustained activation.

Frequencies of CD21^lo^ Bmem are increased up to 28 days post-antigen exposure, and subsequently decrease.^63^ Whilst in both cohorts, the majority of WH1 RBD-specific Bmem were CD21^+^ after both doses, the frequencies of CD21^lo^ WH1 RBD-specific Bmem were increased in controls after dose 3 (**Fig 4 F&G**). This was not associated with increased numbers or frequencies of total CD21^lo^ Bmem (**Supplementary Fig 5 F&G**).

Finally, expression of CD27 is associated with secondary responses for IgG^+^ Bmem.^64^ The majority of RBD-specific IgG^+^ Bmem in controls expressed CD27, and these were significantly higher than in IEI patients at both doses (**Fig 4 H&I**). Also, within total IgG^+^ Bmem, IEI patients also showed reduced CD27 expression (median of 12% vs 90% CD27^+^ Bmem in controls) indicating that this reduced expression might be associated with their general humoral immune memory defect (**Supplementary Fig 5 H**).

### Reduced Ig class switching of WH1 RBD-specific Bmem in IEI patients after third COVID-19 vaccine

To investigate the vaccine-elicited formation of class-switched Bmem in IEI patients, we employed our extensive immunophenotyping including Ig isotype and IgG subclasses (**Fig 5 A**). We previously found expansions of IgG1 after two and three doses of COVID-19 vaccination in controls (**Fig 5 B&C**).^14–16^ Using the same approach, we show that IEI patients have fewer WH1 RBD-specific Bmem that express IgA or any IgG subclass (IgG1, 2, 3 or 4) (**Fig5 B-G**). IgM^+^-only expressing RBD-specific Bmem numbers were similar at post-dose 2 between patients and controls, and tended to be higher in IEI patients than controls at post-dose 3 (**Fig 5B**). Boosting of IgG1^+^ and IgG2^+^ WH1 RBD-specific Bmem numbers was observed in IEI patients after dose 3 but these remained lower than controls (**Fig 5 B-D**). In comparison, expansions of IgG3, IgG1, IgG4 and IgA expressing WH1 RBD-specific Bmem were evident in controls after dose 3, coincident with significantly reduced numbers of unswitched IgD^+^IgM^+^ WH1 RBD-specific Bmem (**Fig 5 B-G**).

**FIG 5.**
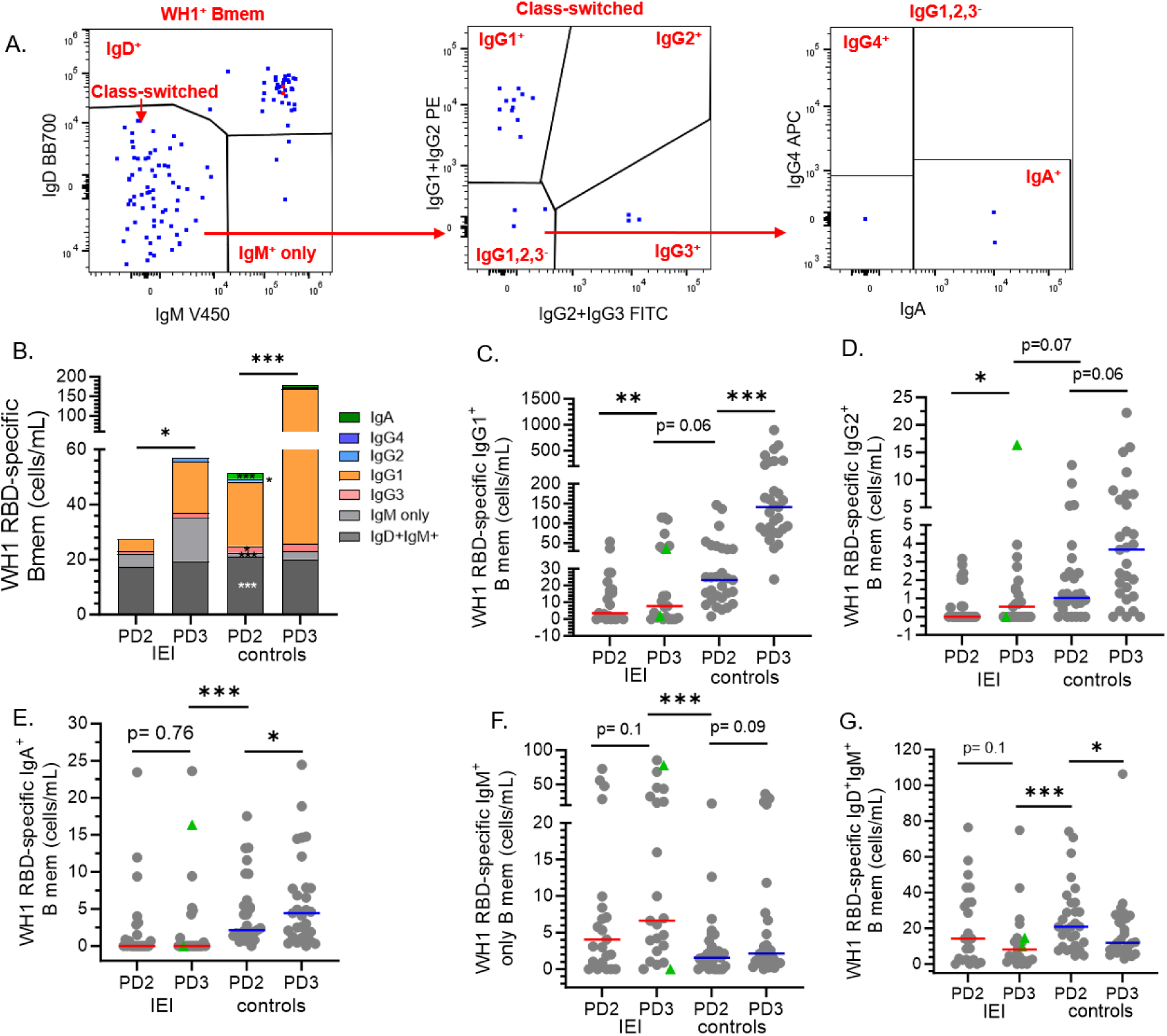
Ig isotype and IgG subclass usage of ancestral RBD-specific Bmem after 2 and 3 COVID-19 vaccine doses in patients with IEI. (**A**) Gating strategy of Ig isotype and IgG subclass expressing RBD-specific Bmem in a representative IEI patient. (**B**) Distribution of Ig isotype and IgG subclass expressing subsets within the ancestral RBD-specific Bmem. Absolute numbers of (**C**) IgG1^+^, (**D**) IgG2^+^, (**E**) IgA^+^, (**F**) IgM^+^ and (**G**) IgD^+^IgM^+^ cells within the WH1 RBD-specific Bmem population after 2 and 3 vaccine doses in all study subjects. Red and blue lines depict medians. Green triangles represent patients with confirmed breakthrough infection (BTI) prior to sampling (**Supplementary Table 3**). Mann-Whitney test was performed for unpaired data and Wilcoxon matched-pairs signed rank test for paired data. *p<0.05, **p<0.001, ***p<0.0001. Healthy donor data previously published in ^14–16^.

Notably, IEI patients had significantly fewer IgG1, IgG2, IgG3 and IgA-class switched, and IgM^+^-only WH1 RBD-specific Bmem than controls (**Fig 5 B-E**). Thus, in addition to reduced vaccine-specific antibody levels (**Fig 1**), IEI patients generate fewer antigen-specific Bmem and these showed reduced maturation after both vaccine doses.

### Third dose vaccine in IEI patients enhances plasma IgG and NAb recognition of Omicron subvariants

The third dose COVID-19 booster vaccine was provided to increase protection against Omicron subvariants, and we previously showed that this dose enhanced NAb, IgG binding and Bmem recognizing Omicron BA.2 and BA.5 in healthy individuals.^15^ In IEI patients, levels of WH1-specific IgG binding to BA.2 and BA.5 RBD were consistently lower than in controls after both doses (**Fig 6 A-C**). Still the third dose significantly increased these levels resulting in higher median levels than healthy controls after dose 2 (**Fig 6 A&B**). None of the IEI patients had detectable NAb to BA.5 after dose 2, and only 5 had detectable NAb after dose 3. In contrast, 76% of controls had detectable NAb to BA.5 after 3 vaccine doses (**Fig 6 C**).

**FIG 6.**
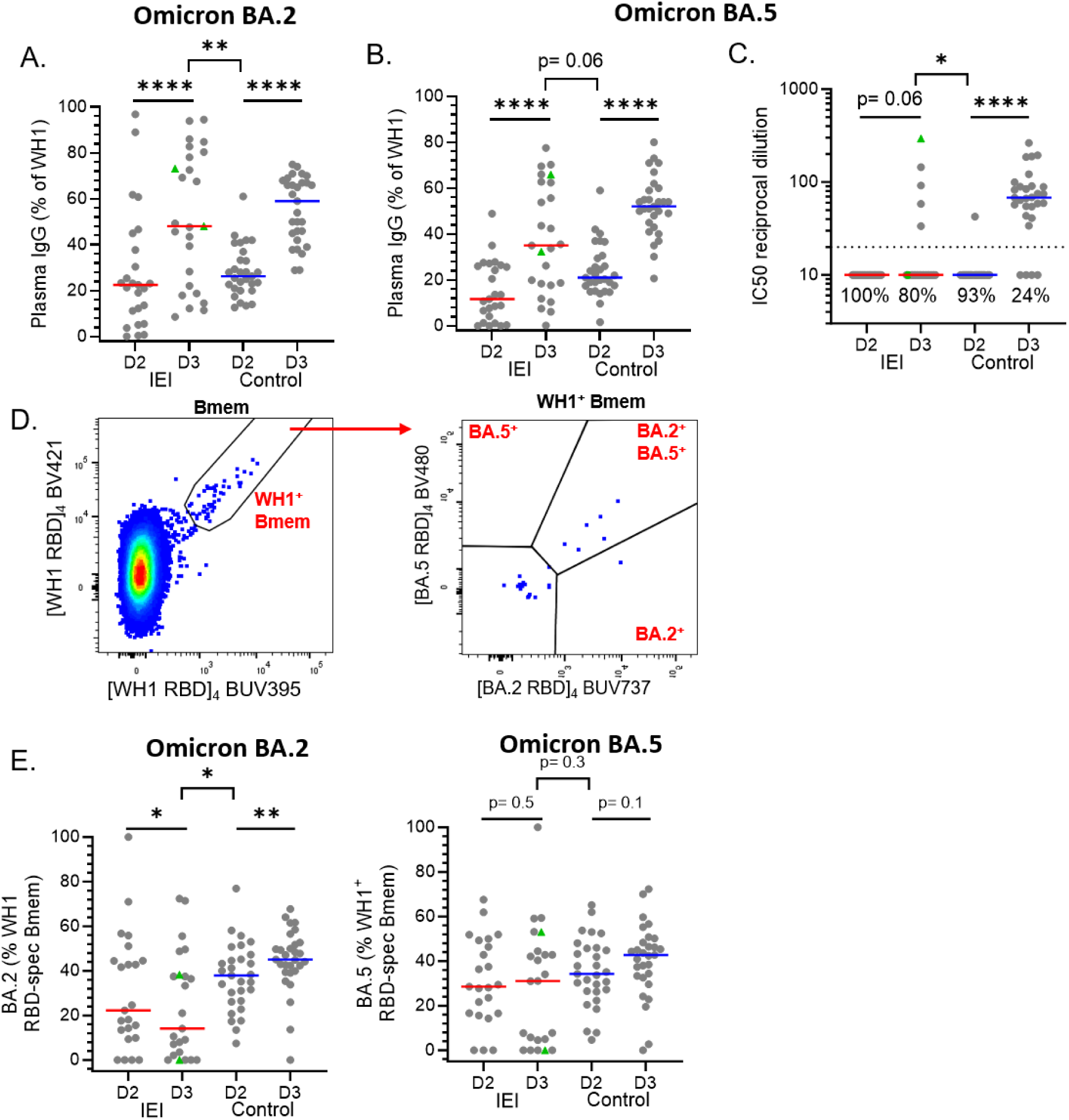
Recognition of Omicron subvariants by WH1 RBD-specific IgG, NAb and Bmem in IEI patients after 2 and 3 COVID19 vaccine doses. Plasma IgG specific for (**A**) Omicron BA.2 and (**B**) BA.5 RBD as a percentage of WH1 RBD-specific IgG. (**C**) NAb against Omicron BA5 SARS-CoV-2. (**D)** Gating strategy detect Omicron BA.2 and BA.5 RBD recognition by ancestral RBD-specific Bmem in representative IEI patient. Frequencies of WH1 RBD-specific Bmem that bound (**E**) Omicron BA2 and/or (**F**) Omicron BA5 RBD. Green triangles represent individuals who had a confirmed breakthrough infection (BTI) prior to sampling (**Supplementary Table 3**). Solid red and blue lines represent median values. Mann-Whitney test was performed for unpaired data and Wilcoxon matched-pairs signed rank test for paired data. *p<0.05, **p<0.001, ***p<0.0001. Healthy donor data previously published in ^14–16^.

The capacity of WH1 RBD-specific Bmem to bind Omicron BA.2 and BA.5 RBDs was examined using fluorescent tetramers of each subvariant (**Fig 6 D**). Within WH1 RBD-specific Bmem from IEI patients, the fractions binding to BA.2 or BA.5 RBDs did not increase after the 3^rd^ dose boost. In contrast, the proportion of WH1 RBD-specific Bmem binding BA2 was significantly increased, with both specificities consistently higher in healthy controls (**Fig 6 E**). These findings indicate that despite the capacity to form immune memory in most IEI patients, this is less efficient than in healthy controls, with reduced capacity for variant recognition upon repeat vaccination.

### Patients who fail to form NAb generate memory B and T cells after dose 3

Significant correlations between ancestral WH1 NAb and IgG levels, and IgG and IgG^+^ Bmem levels were observed in both cohorts (**Supplementary Fig 6 A-B**), whereas significant positive correlation between spike-specific CD4^+^ and CD8^+^ Tmem numbers was only observed for IEI patients (**Supplementary Fig 6 C**). No correlation was observed between the number of spike-specific CD8^+^ or CD4^+^ TFH cells and WH1 RBD-specific IgG or NAb titres (data not shown).

Given that IEI patients with antibody deficiencies may have impaired responses to vaccines,^7, 8, 41, 54, 55, 65, 66^ and that NAbs are the gold standard for assessing vaccine efficacy, we investigated whether those who failed to generate NAb titers could still mount Bmem and/or Tmem responses to COVID-19 vaccination. 4/25 (16%) patients mounted a NAb response after 2 vaccine doses (**Fig 7 A**), compared to 16/25 (64%) after 3 vaccine doses (**Fig 7 B**). Of the 21 patients without an NAb titer, 19/21 (90%) showed a WH1 RBD-specific IgG response equivalent to healthy controls, compared to 4/9 (44%) after dose 3 (**Fig 7 B**). In comparison regardless of dose, all patients with NAb titers had WH1 RBD-specific IgG levels within the healthy range. Together, this infers that whilst dose 3 boosts the number of patients with detectable NAb titres, it does not boost overall WH1 RBD-specific IgG levels in some patients, likely representing a lack of boosting of non-neutralizing responses (**Fig 7 A&B**).

**FIG 7.**
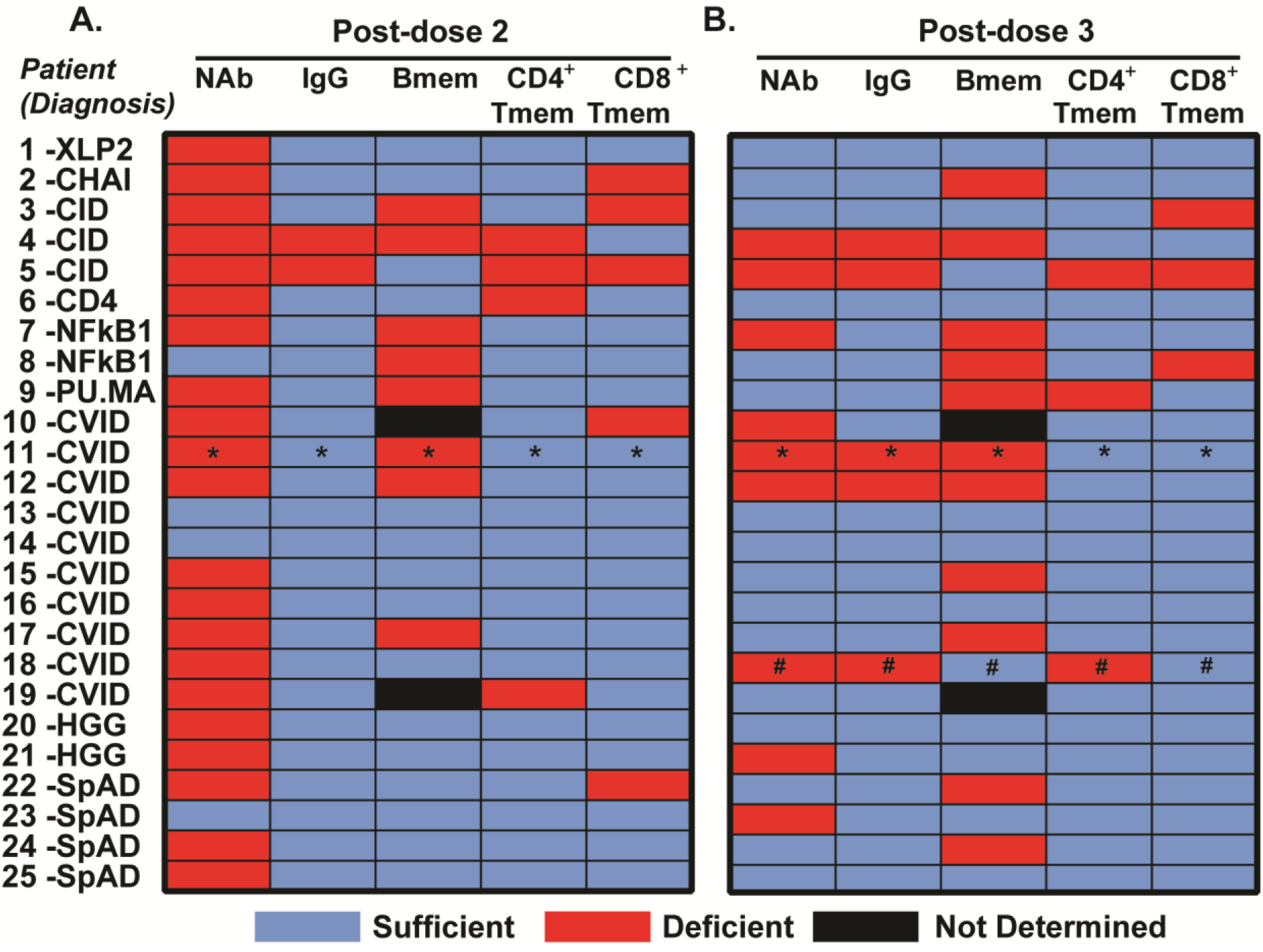
Individual patients can generate Bmem and Tmem, even without detectable NAb titers. Individual patient production of WH1-specific IgG, NAb, Bmem, CD4^+^ and CD8^+^ Tmem (**A**) after dose 2 and (**B**) 3 of monovalent COVID-19 vaccination. Two patients (black) were excluded from Bmem analysis due to low total Bmem. Patients were classified as sufficient (blue) or deficient (red) based on whether they generated NAb titers (>20 IC50 reciprocal dilution) or IgG levels, Bmem, CD4^+^ Tmem or CD8^+^ Tmem numbers within or above the healthy control range. With metrics below these levels deemed deficient. Patients numbered as in **Supplementary Table 1&2**. * blood sampling >100 days after doses 2 and 3. # blood sampling >100 days after dose 3.

After dose 2, all of the 21 patients without an NAb response were able to generate immune memory: 12/19 WH1 RBD-specific Bmem (63%), 17/21 spike-specific CD4^+^ (81%) or 16/21 CD8^+^ (76%) Tmem responses within the healthy range (**Fig 7 A**), which is also demonstrated after dose 3 (**Fig 7 B**). After dose 3, nine patients had no NAb response, including 2 patients with CID, 4 with CVID, 1 with HGG, 1 with NFKB1 haploinsufficiency and 1 with SpAD. Still most of these patients formed spike-specific CD4^+^ Tmem and/or CD8^+^ Tmem (7/9; 78% and 8/9; 89%, respectively) (**Fig 7 B**). Furthermore, of 3/4 (75%) patients that developed NAb titers after dose 2 generated WH1 RBD-specific Bmem and all 4 patients generated CD4^+^ Tmem and CD8^+^ Tmem (**Fig 7 A**). Of the 16 patients that had detectable NAb titres at dose 3, 7/15 generated WH1 RBD-specific Bmem (47%), 15/16 formed CD4^+^ Tmem (94%) and 14/16 CD8^+^ Tmem (88%) (**Fig 7 B**). Thus, all examined patients formed antigen-specific Bmem and/or Tmem in response to COVID-19 vaccination. Of note, three samples were collected more than 100 days after vaccination, and showed no detectable NAb, potentially reflecting natural contraction of the antibody response (**Fig 7**). Here, we show that most IEI patients generate an impaired Ig response but, irrespective of whether NAb titers are generated, all patients form a sufficient immune memory response with respect to Bmem, CD4^+^ Tmem and/or CD8^+^ Tmem. Together this suggests that IEI patients mount some protection after COVID-19 vaccination, however, the degree of protection this affords against severe COVID-19 disease is not yet known. Overall, this demonstrates that regardless of the generation of NAbs all IEI patients had the capacity to generate immune memory in response to vaccination.

## DISCUSSION

We here evaluated the capacity of IEI patients to mount an adaptive immune response to COVID-19 vaccination through comprehensive evaluation of plasma IgG, NAb, Bmem, CD4^+^ and CD8^+^ Tmem specific for WH1 and Omicron subvariants. This study demonstrates that while patients with IEI mount detectable adaptive responses to COVID-19 vaccination, both the magnitude and the quality of these responses are impaired compared to healthy controls. Also, regardless of whether NAb titres are achieved, all IEI patients can generate one or more facets of immune memory e.g. Bmem and/or CD4^+^ Tmem and/or CD8^+^ Tmem in response to COVID-19 vaccination.

Both WH1 and Omicron subvariant IgG levels and NAb titres were markedly reduced after two vaccine doses. The third dose substantially increased seroconversion, but titres remained lower than controls. This is in line with previous studies evaluating NAb and IgG binding levels in IEI patients.^34, 37, 38, 40, 43, 67–69^ Consistent with our previous studies in controls,^15^ levels of WH1 RBD-specific IgG in IEI patients were higher following mRNA than adenoviral vector vaccination at dose 2, and these levels were significantly boosted by a third dose with an mRNA vaccine. This is consistent with other studies where two-dose mRNA vaccination yielded higher IgG levels and NAb titres than those receiving two-dose adenoviral based vaccination.^34, 37, 38, 40, 68, 69^ Here third dose vaccination across both platforms improved seroconversion rates, including in prior non-responders to dose 2.^43, 67, 70^ 13% of the controls in this study did not have NAb after dose 2, which was higher than we previously reported ^15^. This is likely due to the difference in methodology as a gold-standard live neutralization assay was used here as opposed to pseudovirus based assays performed previously. This demonstrates the need for consistent assay use and internal serological controls (e.g. World Health Organization National Institute for Biological Standards and Controls (WHO NIBSC) standard control) when measuring and comparing NAb titers across platforms and studies.^8^

Both Bmem and Tmem are key indicators of immune memory, and represent a better indicator of long-term protection than antibodies which have been shown to wane within 6 months of vaccination.^14–16^ Whilst consistent with numerous studies, we show in this study that IEI patients have reduced numbers of class-switched Bmem in the total Bmem compartment.^54, 55, 64, 71^ For the first time, we extend these investigations to examine activation, maturation and activation of virus-specific Bmem generated to COVID-19 vaccination. Bmem were detectable in most patients with numbers significantly lower than controls after both doses, probably as a consequence of the genetic variants driving IEIs and their role in immune cell function. Therefore, comparing fold changes in Bmem numbers between dose 2 and dose 3 in each cohort, showed that whilst responses were boosted in both cohorts at post-dose 3, this was to a much lower degree in patients than in controls. This is consistent with other studies of IEI patients showing reduced IFNy^+^ B cell generation to COVID-19 vaccination,^36, 38, 51^ as well as reduced Bmem induced to influenza vaccination.^54^

We here observed that IEI patients showed higher frequencies of RBD-specific Bmem expressing CD71 at 4 weeks after vaccination. CD71 was previously found to be expressed on antigen-specific Bmem early after influenza or COVID-19 vaccination (7-14 days), and has been shown to decrease after this point.^14, 16, 72^ The increased CD71 positivity on IEI patient Bmem,, indicates persistent activation and prolonged germinal centre reactions, whereas in controls Bmem were in a quiescent state as evidenced by lower CD71 positivity. Furthermore, we observe reduced IgG subclass and CD27 expression on WH1 RBD-specific Bmem which is indicative of impaired Ig class switching and defective maturation thus translating to impaired formation of long-lived Bmem, the goal of effective vaccination. Whilst detailed studies of IEI after COVID-19 vaccination are lacking, we previously found reduced IgG usage in antibody-deficient patients after influenza booster vaccination. Here, we observed increased CD27 positivity of influenza-specific Bmem,^54^ which is in contrast to CD27 expression shown in this study. These differences could be due to genetic heterogeneity of the study population, the different vaccine antigens used e.g. influenza vs. SARS-CoV-2, and/or the novel COVID-19 vaccine platforms utilised here.

Within the WH1 RBD-specific Bmem, we did not detect differences in the frequencies of CD21^lo^ Bmem between patients and controls. study groups, suggesting that numbers of circulating cells are not associated with poor vaccination responses. A subgroup of CVID patients carry increased frequencies of CD21^lo^ B cells in blood, likely due to a decreased absolute number of naïve B cells.^55, 73, 74^ However, assessment of frequencies might indicate this but were not examined in this context. Therefore, more in depth analysis of the pre-existing Bmem profile both numerically and proportionally may provide markers predicting poor vaccination responses, but this was outside the scope of this study.

All patients in our study formed Spike-specific Tmem, with numbers of spike-specific CD8^+^ Tmem but not CD4^+^ Tmem lower in IEI patients than controls. This is in line with previous studies reporting lower frequencies of CD4^+^ and/or CD8^+^ T cells to COVID-19 vaccination than controls.^39, 42–49^ Our extended evaluation of Tmem showed that spike-specific CD4^+^ or CD8^+^ Tmem have reduced CD69 expression suggesting defects in activation,^75^ which likely influences the defective generation of IFNγ^+^TNFα^+^ Tmem, cells known to strongly associate with elimination of virus-infected cells, and which have long been a focus for improved vaccine efficacy.^76, 77^ Whilst not a focus in this study, a multicentre study of IEI patients has documented that COVID-19 adenoviral vaccination elicited stronger T-cell responses than mRNA vaccination. ^68^ However, analysis in our cohort shows the opposite with numbers of both spike-specific CD4^+^ and CD8^+^ Tmem trending to be decreased in both cohorts receiving the heterologous regimen (2-dose adenovirus plus 1 dose mRNA) compared to the homologous (3 dose mRNA) regimen. Importantly, even in the absence of NAbs, most patients generated spike-specific Bmem and Tmem, indicating that immune memory is formed despite quantitative deficiencies. However, these memory cells showed qualitative impairments, including reduced Ig class-switching of Bmem, diminished Tfh induction, and lower T-cell activation and polyfunctionality. Together, this suggests that although immune memory can be elicited in COVID-19 vaccinated IEI patients, deficits in cellular maturation may compromise breadth and durability of protective immunity.

The heterogeneity in vaccine responses within our cohort underscores how genetic and clinical factors influence vaccine responses: these were most impaired in patients with CVID and CID, while some patients with SpAD or hypogammaglobulinemia achieved NAb, IgG, Bmem and Tmem levels approaching those of controls. Notably, patients with defined monogenic defects such as NFKB1 haploinsufficiency or CHAI failed to generate humoral responses, but formed T-cell memory, highlighting divergence between immune compartments. XLP2 patients demonstrated seroconversion alongside generation of both Bmem and Tmem after dose three vaccination. These findings suggest that vaccine responsiveness cannot be reliably predicted by clinical or genetic phenotype alone, though larger IEI cohorts are needed to confirm this.

It is important to note that age, certain genetic defects, and latent viral infections including EBV and CMV can skew the T-cell compartment toward terminal differentiation and exhaustion, potentially limiting functional memory formation particularly for CD8^+^ T-cells.^65, 78, 79^ Therefore, the interplay between individual genetic defects and environmental exposures such as chronic viral infection and aging can profoundly impact vaccine responsiveness in immunocompromised populations.

Prior SARS-CoV-2 infection has also been shown to enhance T-cell immunity in healthy individuals.^80^ However, its impact on immunity in IEI patients is unknown, and no studies have directly compared the immune responses generated or infection outcomes in vaccinated versus unvaccinated IEI populations. Among patients with serological evidence of BTIs, we observed higher numbers of WH1-specific Bmem and Tmem. However, the low frequency of BTIs in this cohort, due to Australia’s strict public health measures precluded robust statistical analysis. Therefore, further analysis is required to determine whether hybrid immunity confers any protective benefit to IEI patients.

In patients receiving IgRT, NAb titers are difficult to interpret as assays cannot discriminate between circulating antibodies generated endogenously from those administered exogenously. Plasma samples analyzed in this study were obtained in 2021 at the time when SARS-CoV-2 infection and vaccination levels in Australia were low, and where IgRT received would have been donated a minimum of 9 months prior. Thus, it is unlikely that the IgRT products contained SARS-CoV-2 antibodies. Additionally, no patients received SARS-CoV-2 Ab therapies (eg. Evusheld and Sotrovimab) and, therefore, those antibodies detected in patients are *bona fide* responses to vaccination. Here, the use of tetramer-based flow cytometry and AIM assays enabled sensitive detection of vaccine-specific Bmem and Tmem, offering more a comprehensive picture of immune memory than examining circulating antibodies alone. Together, these results underscore the added value of detailed Bmem and Tmem profiling to assess immunity to COVID-19, in addition to quantitating NAb and IgG responses which are the accepted metrics of protection.^9, 31, 34, 35, 81, 82^ This is particularly important in patients receiving IgRT where serum IgG and NAb measurement alone could overestimate vaccine benefits while vaccine-induced antibodies cannot be discriminated from those received in the IgRT. Our findings support a more comprehensive approach to immune monitoring, incorporating antigen-specific Bmem and Tmem alongside antibody measurements. Developing high-throughput assays particularly for Tmem analysis should be a priority, given their value in understanding the vaccine responses of immunodeficient and other at-risk populations.^34, 35^ This may support more accurate risk stratification and guide decisions on booster timing or adjunctive therapies for treatment of infection. Moreover, the impaired recognition of Omicron subvariants observed in our cohort suggests that repeated vaccination with updated formulations may be required to boost responses and counter viral evolution, as reduced breadth of immune memory could prolong infection or increase risk of severe disease in these populations.

This study has several limitations, including a modest cohort size, large clinical and genetic heterogeneity, and the lack of longitudinal follow-up to monitor durability of the immune responses and their correlation with clinical outcomes of breakthrough infections. Mechanistically, further research is needed to assess non-neutralizing antibody functions, cytotoxic T-cell function, and the durability of responses across vaccine platforms, as well as whether additional doses of vaccine improve the quantity and quality of the induced responses. A further point of interest is whether as shown in healthy individuals extending mRNA vaccine dose interval, may enhance SARS-CoV-2-specific CD4^+^ T cell functionality.^83^ Because T-cell help is crucial for generating and sustaining Bmem, longer intervals could also strengthen humoral immunity in some patients. Further analysis is needed to determine where additional doses should be included in primary vaccine regimens for other pathogens (e.g. influenza) and whether patients with failed vaccine responses should receive an extra dose.

Nonetheless, our findings demonstrate that although IEI patients show impaired vaccine responses, the majority can generate measurable immune memory, suggesting that vaccination is likely to confer some degree of protection. These findings underscore the importance of continued prioritization of IEI patients for vaccination, need for repeated vaccination to enhance responses, as well as comprehensive immune monitoring to guide clinical management and public health decisions in vulnerable populations.

## Data Availability

All data produced in the present work are contained in the manuscript.

## ABBREVIATIONS

CVID: common variable immunodeficiency;
HGG: hypogammaglobulinemia;
SpAD: specific antibody deficiency;
XLA: X-linked agammaglobulinemia,
IgRT: immunoglobulin replacement therapy;
PBS: phosphate buffered saline;
Nab: Neutralizing antibody;
IgG: immunoglobulin G;
Bmem: memory B cell;
Tmem: memory T cell.

## Acknowledgements

We are grateful to Mr Jack Edwards, Dr Paul Gill and Ms Alina Wang for technical support, the Alfred pathology collection service and the Alfred Medical Day unit for blood sample collection, Dr Priscilla Auyeung for collation of clinical data, and the ARA flow core facility staff for flow cytometry support. The work was supported by the Australian Medical Research Future Fund (MRFF, project no. 2016108), The Jeffrey Modell Foundation, and an Allergy and Immunology Foundation of Australasia (AIFA) Primary Immunodeficiency Clinical Research Grant (supported by CSL Behring, Australia). RG thanks the Burnet Institute for supporting a sabbatical at Monash University.

## Author Contributions

Designed and/or performed experiments: ESJE, RG, NV, SS, JC, EGB, PMA, IB, HD, MCvZ; Formal analysis: ESJE, RG, and IB; Provided patient care and clinical data: SO, JJB, and SS; Provided reagents and methodology: SJB, ST, AA; Supervised the work: ESJE, RG, and MCvZ; Wrote the manuscript: ESJE, RG and MCvZ. All authors edited and approved the final version of the manuscript.

## Contribution to the field (200 words max)

Patients with inborn errors of immunity (IEI) are at increased risk of severe COVID-19 disease and often exhibit impaired vaccine responses, particularly in the context of antibody deficiency. Most prior studies have focused narrowly on seroconversion as a read-out for vaccine efficacy, but this is complicated by immunoglobulin replacement therapy where donated and recipient antibodies cannot be distinguished between. This leaves major gaps in our understanding of the breadth and quality of immune memory generated to vaccination in this population. In this study, we provide a comprehensive analysis of humoral and cellular responses after two and three doses of COVID-19 vaccination in adults with IEI, encompassing IgG, NAb, memory B cells and both CD4^+^ and CD8^+^ memory T cells. We show that while NAb responses are impaired and variant recognition is reduced, the majority of IEI patients mount detectable Bmem and Tmem responses, even in the absence of NAb titres. This demonstrates the value of multidimensional immune profiling in at-risk populations, and underscores the likely benefit of repeated vaccination. Thus, providing mechanistic insights to guide immunization strategies, monitoring, and therapeutic interventions in IEI patients.

