## Supplementary Data for "Third dose COVID-19 vaccination elicits immune memory in patients with inborn errors of immunity even in absence of neutralizing antibodies"

**SUPPLEMENTARY MATERIALS**

### Supplementary Tables (n=6)

### Supplementary Table 1. Clinical and immunological details of all patients

| Pt | Age at inclusion (yr) | Sex | Diagnosis | Serum Ig (g/L) | | | Immune cells (cells per µl blood) | | | Systemic treatment during study | | Vaccine received | |
|---|---|---|---|---|---|---|---|---|---|---|---|---|---|
| | | | | IgG | IgA | IgM | B cells | CD4+ T cells | CD8+ T cells | IgRT | Other systemic medications | Dose 1+ 2 | Dose 3 |
| **1** | 20-31 | M | XLP2[1] | 14.5 | 6.1 | 3.2 | 275 | 916 | 500 | — | infliximab | BNT162b2 | BNT162b2 |
| **2** | 51-60 | F | CHAI[2] | 10.1 | 3.1 | 0.5 | 142 | 511 | **190** | — | — | ChAdOx1 | BNT162b2 |
| **3** | 21-30 | M | CID | **6.1** | **0.6** | 0.4 | 121 | **221** | **30** | Y | antifungal, mycophenolate | BNT162b2 | BNT162b2 |
| **4** | 21-30 | F | CID | **1.1** | **<0.1** | **<0.1** | 120 | **226** | **104** | Y | — | BNT162b2 | BNT162b2 |
| **5** | 41-50 | M | CID | 10.5 | 1.6 | 1.6 | 293 | **76** | **67** | — | — | BNT162b2 | mRNA-1273 |
| **6** | 51-60 | F | CD4 cytopenia | 11 | 2.2 | 0.5 | 187 | **98** | **182** | — | cotrimoxazole prophylaxis | ChAdOx1 | BNT162b2 |
| **7** | 21-30 | F | PU.MA[3] | **1.1** | **0.4** | **0.2** | 166 | **232** | **93** | Y | prednisolone, budesonide | BNT162b2 | BNT162b2 |
| **8** | 21-30 | F | NFKB1 haplo | **6.9** | **<0.1** | **0.2** | 174 | **218** | 273 | Y | rituximab | BNT162b2 | BNT162b2 |
| **9** | 41-50 | F | NFKB1 haplo | **6.8** | 2.1 | 0.5 | 239 | 799 | 426 | Y | doxycycline, prednisolone intermittently, mycophenolate | BNT162b2 | BNT162b2 |
| **10** | 51-60 | M | CVID | **4.5** | 2 | **<0.1** | **59** | 605 | **55** | Y | lisdexamfetamine | BNT162b2 | BNT162b2 |
| **11** | 51-60 | F | CVID | **6.7** | **<0.3** | **<0.2** | 273 | 807 | **122** | Y | dehydroepiandrosterone, escitalopram, rabeprazole, meloxicam, loratadine | ChAdOx1 | mRNA-1273 |
| **12** | 61-70 | M | CVID | 12.3 | **<0.1** | **<0.1** | 144 | 387 | 676 | Y | — | ChAdOx1 | BNT162b2 |
| **13** | 21-30 | F | CVID | **4.4** | **0.2** | 0.4 | 109 | 668 | 560 | Y | — | BNT162b2 | BNT162b2 |
| **14** | 31-40 | F | CVID | **5.2** | **0.3** | 0.6 | 117 | 1204 | 428 | Y | — | BNT162b2 | BNT162b2 |
| **15** | 31-40 | F | CVID | **0.4** | **<0.1** | **<0.1** | 199 | 576 | 516 | Y | — | BNT162b2 | BNT162b2 |
| **16** | 31-40 | F | CVID | **2** | **<0.1** | **<0.1** | 252 | 530 | **208** | Y | — | BNT162b2 | BNT162b2 |
| **17** | 41-50 | F | CVID | **1.4** | **<0.1** | 1.1 | **50** | **226** | 236 | Y | desvenlafaxine | BNT162b2 | BNT162b2 |
| **18** | 51-50 | F | CVID | **1.7** | **<0.1** | **0.1** | 271 | 644 | 334 | Y | — | BNT162b2 | mRNA-1273 |
| **19** | 51-60 | F | CVID | **6.9** | **0.1** | **0.3** | 106 | 802 | **193** | — | — | ChAdOx1 | BNT162b2 |
| **20** | 21-30 | F | HGG | **4.3** | 1.3 | 0.7 | 352 | 1235 | 860 | Y | seroquel | BNT162b2 | BNT162b2 |
| **21** | 41-50 | F | HGG | **4.7** | 1.4 | 0.9 | 112 | 527 | 339 | Y | hormone replacement therapy | BNT162b2 | BNT162b2 |
| **22** | 41-50 | F | SpAD | **6.6** | 1.1 | 0.7 | 190 | 1142 | **207** | Y | sitagliptin/metformin, rosuvastatin, pantoprazole, amitriptyline, fluticasone propionate/salmeterol, doxycycline | BNT162b2 | BNT162b2 |
| **23** | 51-60 | F | SpAD | 9.3 | 2.2 | 0.8 | 115 | 509 | 386 | — | rabeprazole | BNT162b2 | BNT162b2 |
| **24** | 51-60 | M | SpAD | 10.5 | 1.1 | **0.2** | **37** | 938 | 282 | Y | — | ChAdOx1 | BNT162b2 |

| Pt | Age at inclusion (yr) | Sex | Diagnosis | Serum Ig (g/L) | | | Immune cells (cells per µl blood) | | | Systemic treatment during study | | Vaccine received | |
|---|---|---|---|---|---|---|---|---|---|---|---|---|---|
| | | | | IgG | IgA | IgM | B cells | CD4+ T cells | CD8+ T cells | IgRT | Other systemic medications | Dose 1+ 2 | Dose 3 |
| **25** | 61-70 | M | SpAD | **5.3** | **0.7** | 0.9 | 352 | 613 | 518 | Y | — | ChAdOx1 | BNT162b2 |

Values below normal range are depicted in **bold** font and above are underlined. Diagnostic laboratory Ig reference ranges: IgG, 7.5-15.6 g/L; IgA, 0.85-4.99 g/L; IgM, 0.35-2.42 g/L. 5$^{th}$-95$^{th}$ percentiles of immune cells: B cells, 97-614 cells/µl; CD4$^{+}$ T cells, 319-1475 cells/µl; CD8$^{+}$ T cells, 222-989 cells/µl.[4]
CHAI, Cytotoxic T lymphocyte antigen 4 haploinsufficiency with autoimmune infiltration; CID, combined humoral and cellular immunodeficiency; CVID, common variable immunodeficiency; HGG, hypogammaglobulinemia; IgRT, immunoglobulin replacement therapy; NFKB1 haplo, nuclear factor kappa-B subunit 1 haploinsufficiency; PU.MA, PU.1-mutated agammaglobulinemia; SpAD, specific antibody deficiency; XLP-2, X-linked lymphoproliferative syndrome-2.

**Supplementary Table 2. Participant characteristics**

| | All | | | Heterologous | | | Homologous | | |
|---|---|---|---|---|---|---|---|---|---|
| | **Control** (n=29) | **IEI** (n=25) | ***p*-value**[1] | **Control** (n=15) | **IEI** (n=7) | ***p*-value**[1] | **Control** (n=14) | **IEI** (n=18) | ***p*-value**[1] |
| *Interval between doses (days between-doses; median and range)* | | | | | | | | | |
| dose 1 to dose 2 | 78 (21-88) | 28 (20-97) | 0.29 | 84 (78-88) | 84 (70-86) | >0.99 | 24 (21-39) | 24 (20-97) | 0.71 |
| dose 2 to dose 3 | 188 (154-310) | 112 (57-199) | **<0.0001** | 184 (154-202) | 112 (56-128) | **<0.0001** | 198 (186-310) | 109 (57-199) | **<0.0001** |
| *Timing of blood sampling (days post-vaccination; median and range)* | | | | | | | | | |
| 1-month post-dose 2 | 28 (25-35) | 30 (20-148) | 0.12 | 28 (25-35) | 31 (27-39) | 0.17 | 28 (27-32) | 30 (20-148) | 0.32 |
| 1-month post-dose 3 | 29 (27-43) | 30 (25-169) | 0.57 | 29 (27-43) | 31 (27-169) | 0.85 | 31 (27-34) | 32 (25-107) | 0.78 |
| Age (years; median w range) | 38 (24-62) | 48 (23-66) | 0.23 | 37 (29-61) | 58 (52-66) | **0.0003** | 38 (24-62) | 38 (23-58) | 0.86 |
| % Female | 72% (21/29) | 72% (18/25) | >0.99[2] | 87% (13/15) | 57% (4/7) | 0.27[2] | 57% (8/14) | 78% (14/18) | 0.26[2] |
| BNT162b2 at dose 3 | 97% (28/29) | 88% (22/25) | 0.33[2] | 93% (14/15) | 86% (6/7) | >0.99[2] | 100% (14/14) | 89% (16/18) | 0.49[2] |

[1] Non-parametric Mann-Whitney test with Bonferroni correction for multiple comparisons. [2] Chi-square test

**Supplementary Table 3. Comorbidities and outcomes of breakthrough infection**

| Pt | Clinical Diagnosis | Infectious complications | Non-infectious complications | Confirmed SARS-CoV-2 infection pre-dose 3 | Severity of COVID-19 | COVID-19 Treatment |
|---|---|---|---|---|---|---|
| **1** | XLP2 [1] | presumed chronic osteomyelitis | inflammatory bowel disease, cystic acne, pyoderma gangrenosum | N | - | - |
| **2** | CHAI [2] | mild sinopulmonary infections | transient colitis, alopecia universalis, asthma, allergic rhinitis, autoimmune gastritis/pernicious anaemia | N | - | - |
| **3** | CID | pulmonary aspergillosis, mild bronchiectasis | GLILD, AR, eczema, acne, synovitis, bony infarcts | N | - | - |
| **4** | CID | recurrent sinopulmonary infections, CIN III, mild bronchiectasis, thrush | relapsing remitting LAD, suspected GLILD, coeliac, ITP, headaches, mild neutropenia, splenomegaly | N | - | - |
| **5** | CID | multifocal tuberculosis | AR, eczema, food allergies, Autoimmune thyroid disease | N | - | - |
| **6** | CD4 cytopenia | None | None | N | - | - |
| **7** | NFKB1 haplo | tonsillitis, otitis media, sinopulmonary, VZV | ITP, lymphadenopathy, coeliac, migraines, asthma | N | - | - |
| **8** | NFKB1 haplo | None | autoimmune liver disease (unclassified), fibromyalgia, rosacea, asthma, inflammatory arthropathy | N | - | - |
| **9** | PU.MA[3] | HSV, warts, thrush, sinopulmonary | autoimmune gastritis, hepatitis, exocrine pancreatic insufficiency, hidradenitis suppurative, thrombocytopenia, psoriasis | N | - | - |
| **10** | CVID | recurrent sinopulmonary infections | None | N | - | - |
| **11** | CVID | otitis media, sinusitis, pneumonia | None | N | - | - |
| **12** | CVID | bronchiectasis with recurrent sinopulmonary infections, | None | N | - | - |
| **13** | CVID | otitis media, sinusitis | enteropathy (duodenal/coeliac) | N | - | - |
| **14** | CVID | sinusitis, tonsillitis, bronchitis, pyelonephritis | asthma/COPD | N | - | - |
| **15** | CVID | recurrent sinopulmonary infections, gastroenteritis | psoriasis, acne, splenomegaly | N | - | - |
| **16** | CVID | sinusitis, giardia | asthma/COPD, enteropathy | N | - | - |
| **17** | CVID | sinusitis, pneumonia, oral HSV | lichen planus, pernicious anaemia, urticaria, breast cancer | N | - | - |
| **18** | CVID | sinusitis, pneumonia, giardia | splenomegaly, inflammatory bowel disease | Y | Mild | Paxlovid |

| Pt | Clinical Diagnosis | Infectious complications | Non-infectious complications | Confirmed SARS-CoV-2 infection pre-dose 3 | Severity of COVID-19 | COVID-19 Treatment |
|---|---|---|---|---|---|---|
| **19** | CVID | recurrent sinopulmonary infections, bronchiectasis, pulmonary nocardia | low platelets, positive anca with high titre MPO (no clear clinical disease) | N | - | |
| **20** | HGG | recurrent sinopulmonary infections, viral gastroenteritis, recurrent tonsillitis, recurrent oral HSV, giardiasis, VZV, HPV | Asthma | N | - | - |
| **21** | HGG | sinusitis, pneumonia | None | N | - | - |
| **22** | SpAD | otitis media, tonsillitis, sinusitis, pneumonia | None | N | - | -- |
| **23** | SpAD | sinusitis | asthma | Y | Mild | paracetamol, ibuprofen |
| **24** | SpAD | recurrent sinopulmonary infections | Graves' disease, psoriasis, neutropenia, atopy | N | - | - |
| **25** | SpAD | sinopulmonary infections | atopy | N | - | - |

AR, allergic rhinitis; CHAI, Cytotoxic T lymphocyte antigen 4 haploinsufficiency with autoimmune infiltration; CID, combined humoral and cellular immunodeficiency; CIN III, cervical intraepithelial neoplasia grade 3 (severe); COPD, chronic obstructive pulmonary disease;CVID, common variable immunodeficiency; GLILD, granulomatous lymphocytic interstitial lung disease; HGG, hypogammaglobulinemia; HPV, human papillomavirus; HSV, herpes simplex virus; ITP, immune thrombocytopenia; LAD, lymphadenopathy; MPO, myeloperoxidase; NFKB1, nuclear factor kappa-B subunit 1; NIC, non-infectious complications; PU.MA, PU.1-mutated agammaglobulinemia; SpAD, specific antibody deficiency; VZV, varicella zoster virus; XLP-2, X-linked lymphoproliferative syndrome-2.

### Supplementary Table 4. Antibody panel compositions for TruCount and T-cell evaluation on the BD FACSLyric

| **Fluorochrome** | BV421 | HorV500 | BV605 | BV650 | BV711 | BV786 | FITC | PerCP Cy-5.5 | PE | PE-Cy7 | APC | APC-Cy7 | R718 |
|---|---|---|---|---|---|---|---|---|---|---|---|---|---|
| **1. Trucount** | — | — | — | — | — | — | CD3 | CD45 | CD16+CD56 | CD4 | CD19 | CD8a | — |
| **2. Tmem** | CXCR5 | CD3 | CCR7 | — | CD45RA | CD4 | IFNγ | TNFα | CD134 | CD69 | CD137 | — | CD8a |

### Supplementary Table 5. Antibody panel composition for B-cell evaluation on the Cytek Aurora

| **Fluorochrome** | BUV 395 | BUV 737 | BUV 805 | BV421 | cFluor V450 | BV480 | BV605 | BV711 | BV786 | FITC | BB700 | PE | PE-Vio615 | cFluor BYG710 | BYG781 | APC | ViaDye Red |
|---|---|---|---|---|---|---|---|---|---|---|---|---|---|---|---|---|---|
| **3. Bmem** | WH1 RBD | BA.2 RBD | CD3 | WH1 RBD | IgM | BA.5 RBD | CD38 | CD21 | CD71 | IgG2+ IgG3 | IgD | IgG1+ IgG2 | IgA | CD19 | CD27 | IgG4 | Fixable viability |
| **4. Strep control** | Strep | Strep | CD3 | Strep | — | Strep | — | — | — | — | IgD | — | — | CD19 | CD27 | — | Fixable viability |

**Supplementary Table 6. Antibody details**

| Marker | Fluoro-chrome | Clone | Vendor | Cat. Number | Volume (µL)/ 100µL test | Tube |
|---|---|---|---|---|---|---|
| CCR7 | BV605 | 2-L1-A | BD Biosciences | 566754 | 5 | 4 |
| CD3 | BUV805 | UCHT1 | BD Biosciences | 612896 | 2.5 | 2, 3 |
| CD3 | FITC | SK7 | BD Biosciences | 662995* | 46ng | 1 |
| CD3 | V500 | UCHT1 | BD Biosciences | 561416 | 5 | 4 |
| CD4 | PE-Cy7 | SK3 | BD Biosciences | 662995* | 30ng | 1 |
| CD4 | BV786 | SK3 | BD Biosciences | 563877 | 1 | 4 |
| CD8a | APC-Cy7 | SK1 | BD Biosciences | 662995* | 126ng | 1 |
| CD8 | R718 | SK1 | BD Biosciences | 567354 | 1 | 4 |
| CD16 | PE | B73.1 | BD Biosciences | 662995* | 33ng | 1 |
| CD19 | APC | SJ25C1 | BD Biosciences | 662995* | 46ng | 1 |
| CD19 | cFluor BYG710 | HIB19 | Cytek Biosciences | SKU-R7-20010 | 1 | 2, 3 |
| CD21 | BV711 | B-ly4 | BD Biosciences | 563163 | 5 | 2 |
| CD27 | cFluor BYG781 | O323 | Cytek Biosciences | Custom | 0.5 | 2, 3 |
| CD38 | BV605 | HB7 | BD Biosciences | 562665 | 0.2 | 2 |
| CD45 | PerCP Cy5.5 | 2D1 | BD Biosciences | 662995* | 120ng | 1 |
| CD45RA | BV711 | 5H9 | BD Biosciences | 740806 | 0.25 | 4 |
| CD56 | PE | NCAM16.2 | BD Biosciences | 662995* | 22ng | 1 |
| CD69 | PE Cy7 | FN50 | BD Biosciences | 557745 | 0.625 | 4 |
| CD71 | BV786 | M-A712 | BD Biosciences | 563768 | 1 | 2 |
| CD134 | PE | ACT35 | BD Biosciences | 555838 | 5 | 4 |
| CD137 | APC | 4B4-1 | BD Biosciences | 550890 | 2.5 | 4 |
| CXCR5 | BV421 | RF8B2 | BD Biosciences | 562747 | 5 | 4 |
| Fixable Viability | ViaDye Red | - | Cytek Biosciences | SKU-R7-60008 | 0.025 | 2, 3 |
| IFN | FITC | B27 | BD Biosciences | 552887 | 1 | 4 |
| IgA | PE-Vio615 | REA1014 | Miltenyi Biotec | 130-116-882 | 1.5 | 2 |
| IgD | BB700 | IA6-2 | BD Biosciences | 566538 | 1 | 2, 3 |
| IgG1 | PE | G17-1 | BD Biosciences | 624049 | 0.1 | 2 |
| IgG2 | FITC | HP6002 | BD Biosciences | 624045 | 0.5 | 2 |
| IgG2 | PE | HP6002 | BD Biosciences | 624049 | 1 | 2 |
| IgG3 | FITC | HP6047 | BD Biosciences | 624045 | 0.5 | 2 |
| IgG4 | APC | SAG4 | Cytognos | CYT-IGG4AP | 2 | 2 |
| IgM | cFluor V450 | MHM88 | Cytek Biosciences | Custom | 0.25 | 2 |
| Streptavidin | BUV395 | - | BD Biosciences | 564176 | 0.67 | 2, 3 |
| Streptavidin | BUV496 | - | BD Biosciences | 612961 | 0.67 | 2, 3 |
| Streptavidin | BUV615 | - | BD Biosciences | 613013 | 0.67 | 2, 3 |
| Streptavidin | BUV737 | - | BD Biosciences | 612775 | 0.67 | 2, 3 |
| Streptavidin | BV421 | - | BioLegend | 405225 | 0.13 | 2, 3 |
| Streptavidin | BV480 | - | BD Biosciences | 564876 | 0.67 | 2, 3 |
| Streptavidin | BV650 | - | BioLegend | 563855 | 0.13 | 2, 3 |
| TNF | PerCP Cy5.5 | Mab11 | BD Biosciences | 560679 | 0.25 | 4 |
| *Antibodies part of the Multitest$^{TM}$ 6-color TBNK kit (BD Biosciences, Cat. number 662967) | | | | | | |

**Supplementary Figures** (n=6)

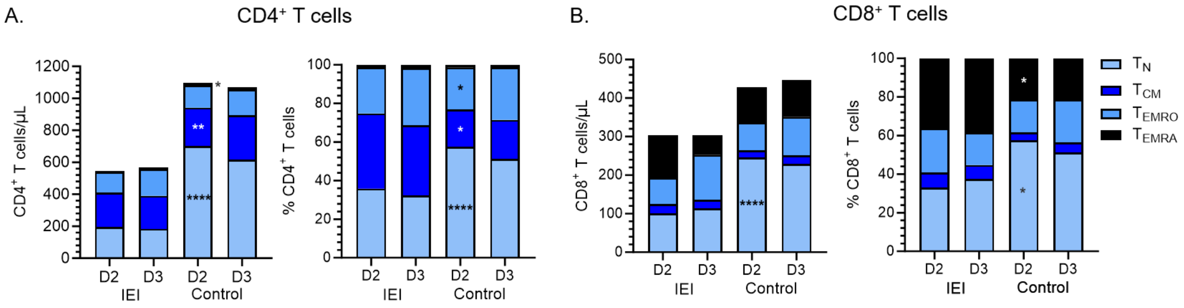


**Supplementary Figure 1: Total $CD4^+$ and $CD8^+$ T cell numbers before and after ancestral COVID-19 vaccine doses 2 and 3 in 25 patients with IEI and 29 healthy controls.** Absolute and relative (**A**) $CD4^+$ and (**B**) $CD8^+$ T cell numbers. Mann-Whitney test was performed for unpaired data and Wilcoxon matched-pairs signed rank test for paired data. Statistical analysis between dose 2 IEI and control is shown by $^{*}p<0.05$, $^{**}p<0.001$, $^{****}p<0.00001$.

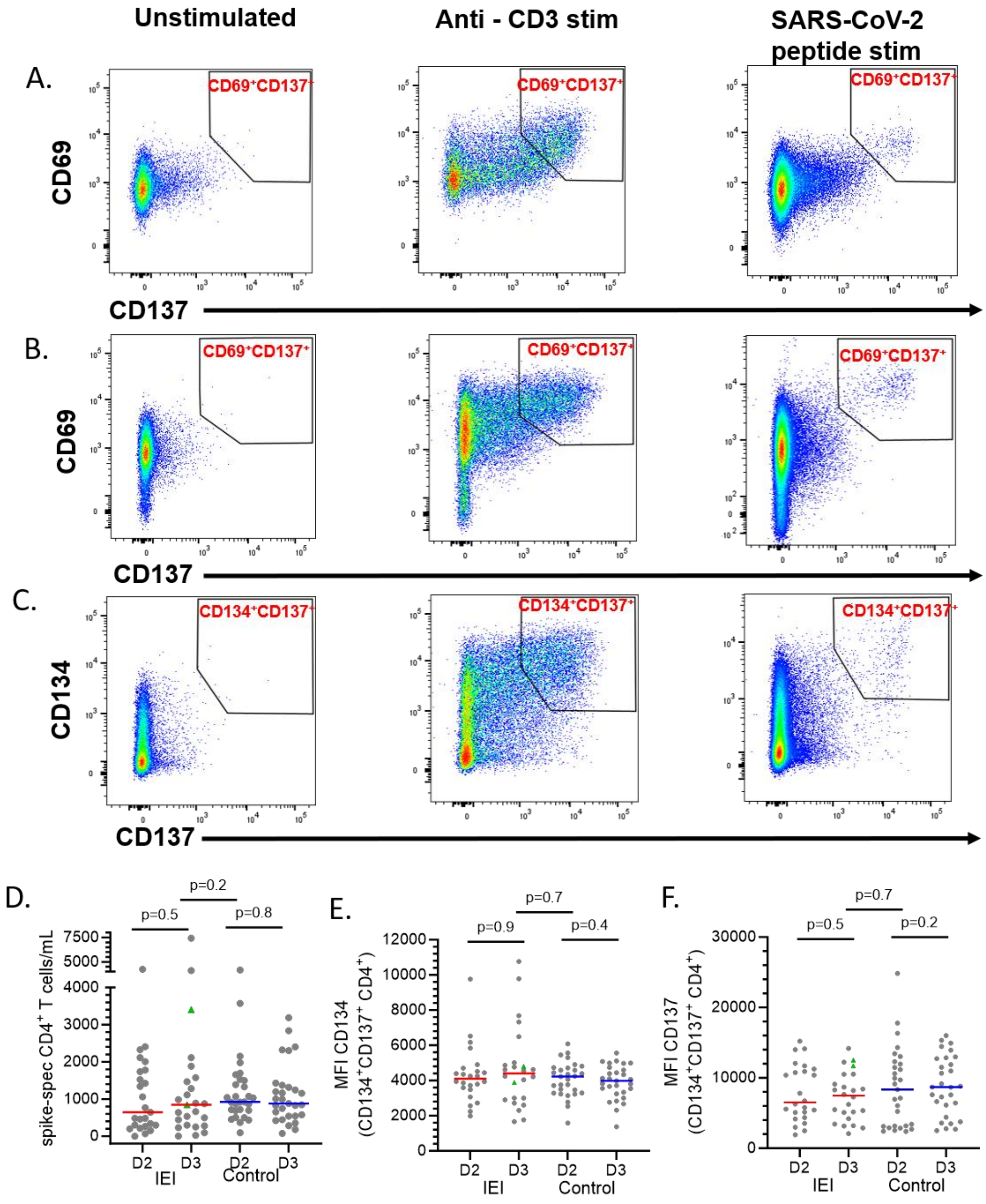


**Supplementary Figure 2: Activation Induced Marker (AIM) co-staining for spike-specific CD4$^+$ or CD8$^+$ T cells.** Intracellular (**A**) CD69 and CD137 in CD8$^+$ T cells, (**B**) CD69 and CD137 expression and (**C**) CD134 and CD137 in CD4$^+$ T cells. (**D**) Absolute spike-specific CD4$^+$ T cell numbers, defined by co-expression of CD134 and CD137. Mean fluorescence intensity (MFI) of (**E**) CD134 and (**F**) CD137 on spike-specific CD4$^+$ T cells. Green triangles indicate individuals with breakthrough infection (BTI) before sampling (**Supplementary Table 6**). Red and blue lines represent median values. Mann-Whitney test for unpaired data and Wilcoxon matched-pairs signed rank test for paired data. *p<0.05, **p<0.001, ***p<0.0001.

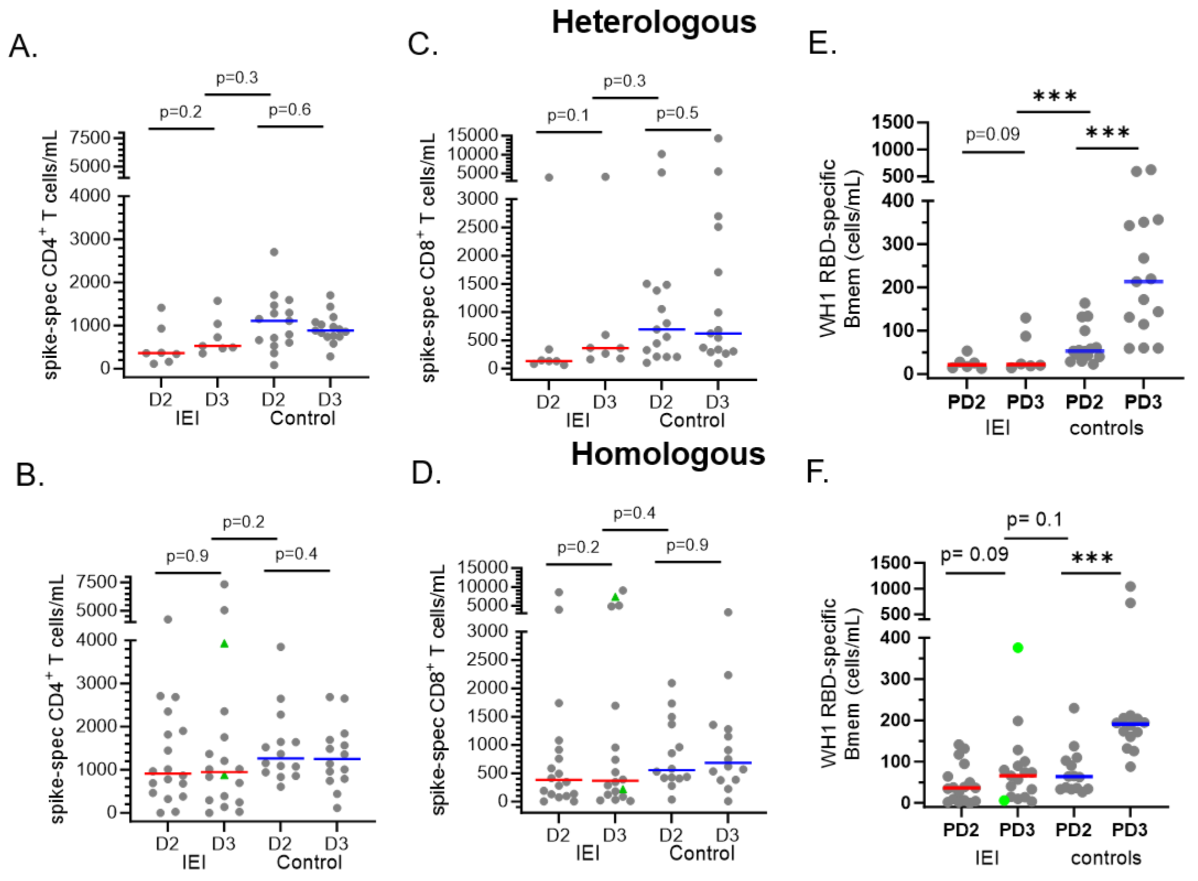


**Supplementary Figure 3: Ancestral CD4+ Tmem, CD8+ Tmem and Bmem after 2 and 3 COVID-19 vaccine doses.** Samples included 1-month post-dose 2 and 1-month post-dose 3 samples from IEI patients and healthy controls receiving primary ChAdOx1 (heterologous) or BNT162b2 (homologous) vaccination followed by a 3rd dose mRNA vaccine. Spike-specific CD4+ Tmem numbers to (**B**) heterologous and (**C**) homologous vaccination. Spike-specific CD8+ Tmem numbers in (**D**) heterologous and (**E**) homologous vaccine recipients, and WH1 RBD-specific Bmem numbers in (**F**) heterologous and (**G**) homologous recipients. Green triangles show confirmed breakthrough infections (BTI) prior to sampling (**Supplementary Table 6**). Red and blue lines represent median values. Mann-Whitney test and Wilcoxon matched-pairs signed rank test used for unpaired and paired data, respectively (*p<0.05, **p<0.001, ***p<0.0001). Healthy donor data previously published in [5, 6, 7].

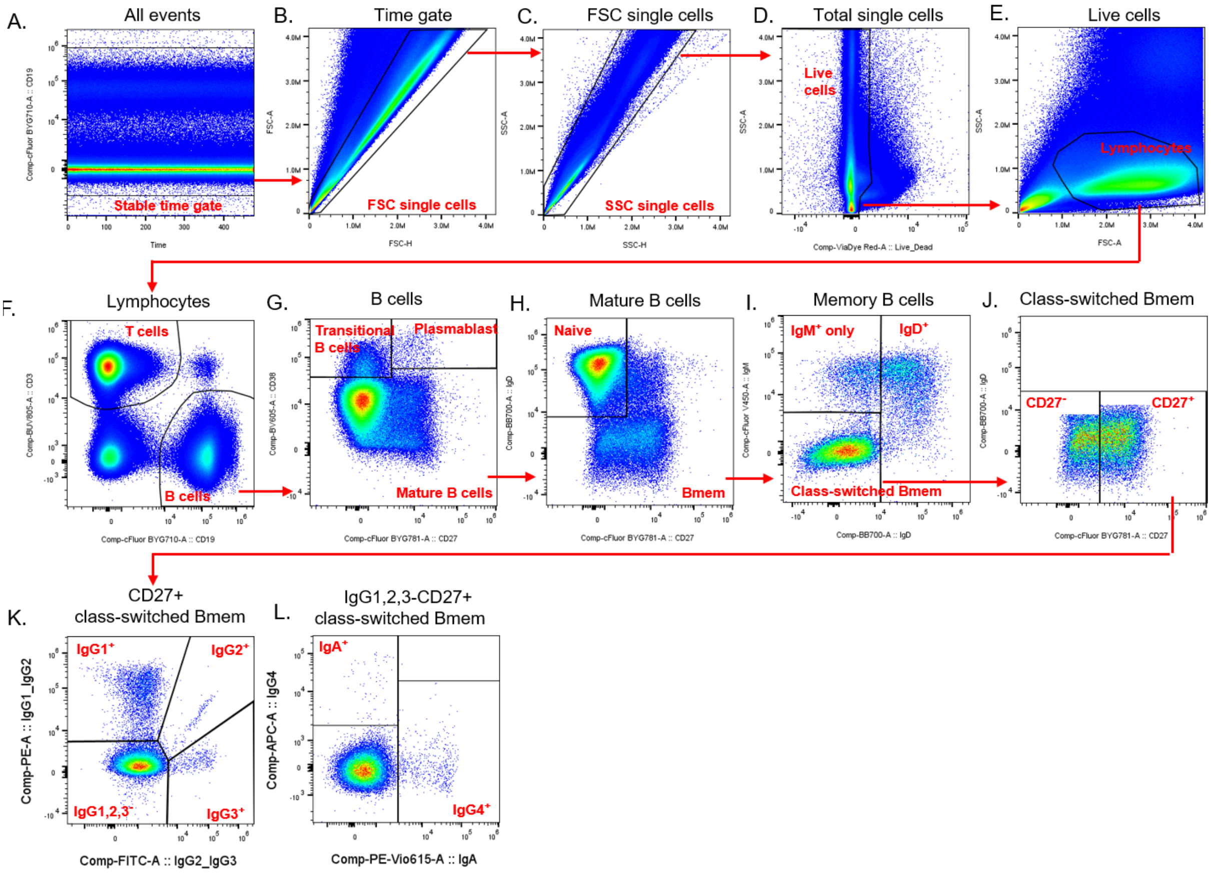


**Supplementary Figure 4: B-cell gating strategy**. (**A**) All events were gated on CD19 vs time to ensure the steady state of the CD19 signal across acquisition time and to exclude aggregates. (**B-C**) Doublets were excluded on FSC-A vs. FSC-H, then SSC-A vs. SSC-H. (**D**) Dead cells were excluded on SSC-A vs. viability dye. (**E**) Lymphocytes were gated as $SSC^{lo}FSC^{mid}$. (**F**) B cells were gated as $CD19^{+}CD3^{-}$. (**G**) Mature, transitional and plasmablasts were gated using CD38 vs. CD27. (**H**) Memory B cells (Bmem) and naive B cells were gated on IgD vs. CD27 in mature B cells. (**I**) Class-switched, $IgD^{+}$ and $IgM^{+}$ only mature Bmem cells were gated on IgM vs. IgD. (**J**) $CD27^{-}$ and $CD27^{+}$ class-switched mature Bmem were gated on CD27 vs IgD. (**K**) $IgG1^{+}$, $IgG2^{+}$, $IgG3^{+}$, and IgG1,2,3- class-switched mature Bmem were gated within $CD27^{+}$ cells (and $CD27^{-}$ cells plot not shown). (**L**) $IgG4^{+}$, $IgA^{+}$ and unclassed mature Bmem were gated within $IgG1,2,3^{-}CD27^{+}$ cells (and $CD27^{-}$ cells plot not shown).

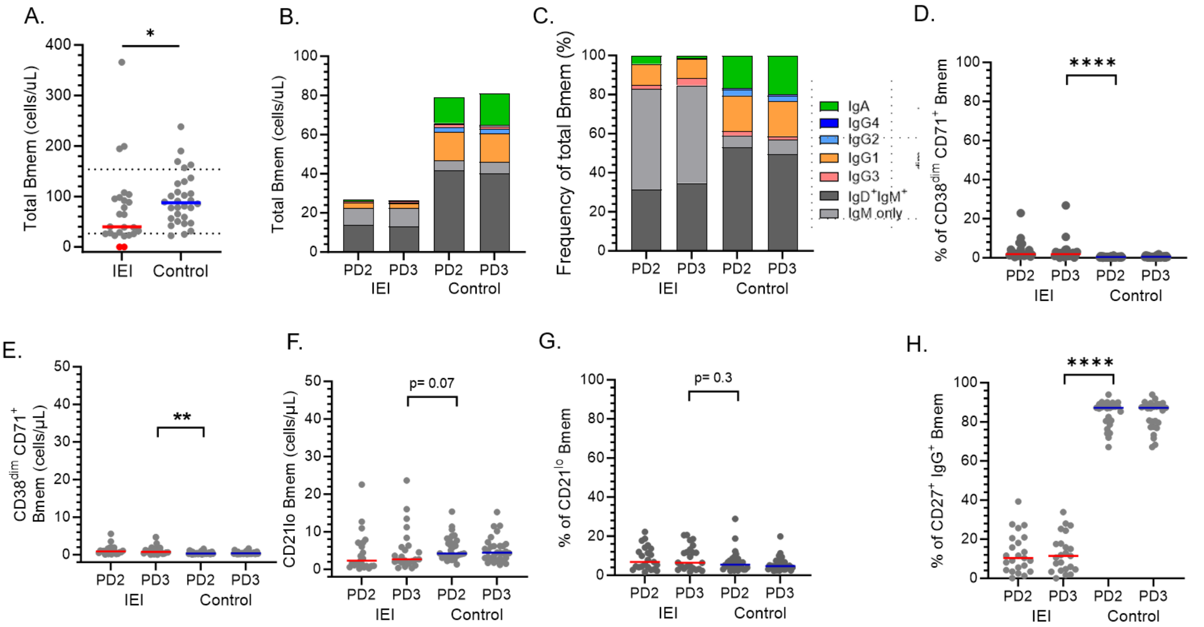


**Supplementary Figure 5: Total memory B cell numbers and IgG isotypes before and after monovalent COVID-19 dose 2 and 3.** (**A**) Absolute numbers of total memory B cells (Bmem) at recruitment. The red dots represent patients with undetectable Bmem, which were excluded from further analyses. (**B**) Absolute numbers and (**C**) frequencies of IgG1$^+$, IgG2$^+$, IgG3$^+$, IgG4$^+$, IgA$^+$, IgM only and IgD$^+$IgM$^+$ cells within total Bmem. (**D**) Absolute numbers and (**E**) frequencies of CD71$^+$. (**F**) Absolute numbers and (**G**) frequencies of CD21$^{lo}$ Bmem cells. (**H**) Frequencies CD27$^+$ within total IgG$^+$ Bmem cells. Green triangles represent individuals who had a confirmed breakthrough infection (BTI) prior to sampling (**Supplementary Table 4**). Red and blue lines in all panels represent median values. Statistics: Mann-Whitney test for unpaired data and Wilcoxon matched-pairs signed rank test for paired data. *p<0.05, **p<0.001, ***p<0.0001. Healthy donor data previously published in [5, 6, 7].

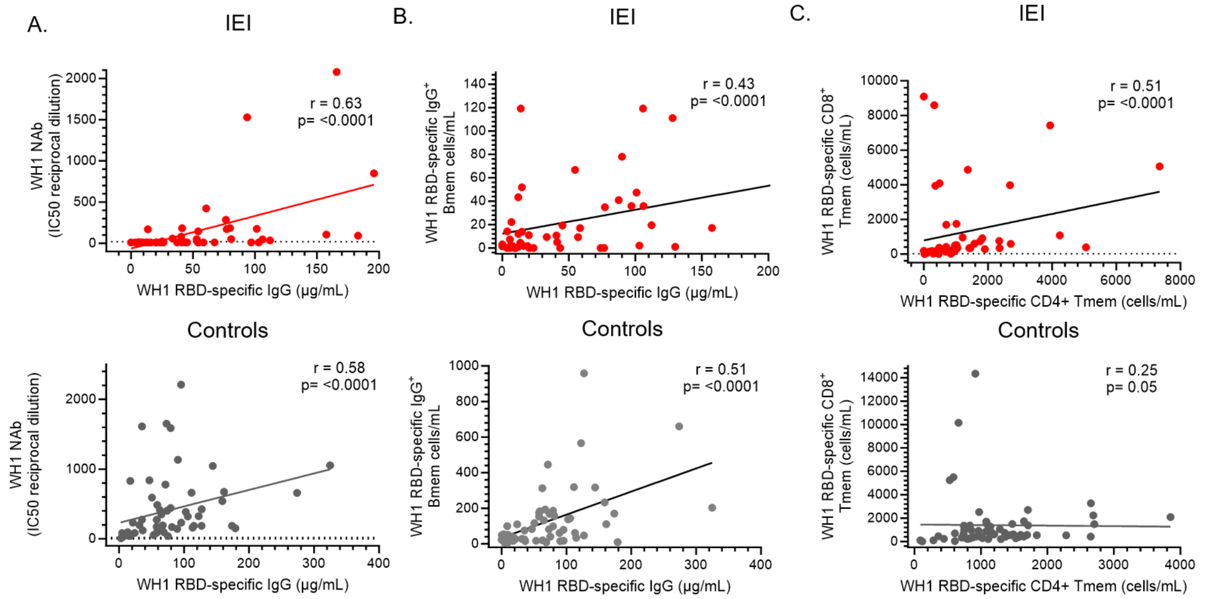


**Supplementary Figure 6: Correlation between markers of humoral and cellular immunity to COVID-19 vaccination.** Correlation between (**A**) WH1 RBD-specific IgG and Nab, (**B**) WH1 RBD-specific IgG and absolute numbers of IgG$^{+}$ WH1 RBD-specific Bmem, (**C**) WH1 spike-specific CD4$^{+}$ Tmem and CD8$^{+}$ Tmem were assessed in IEI patients and healthy controls. Dotted line in (**A**) indicates the neutralization cut-off at an IC50 value of 20. Statistics were performed using Spearman's rank correlation. Healthy donor data previously published in [5, 6, 7].
